# Early introductions and community transmission of SARS-CoV-2 variant B.1.1.7 in the United States

**DOI:** 10.1101/2021.02.10.21251540

**Authors:** Tara Alpert, Anderson F. Brito, Erica Lasek-Nesselquist, Jessica Rothman, Andrew L. Valesano, Matthew J. MacKay, Mary E. Petrone, Mallery I. Breban, Anne E. Watkins, Chantal B.F. Vogels, Chaney C. Kalinich, Simon Dellicour, Alexis Russell, John P. Kelly, Matthew Shudt, Jonathan Plitnick, Erasmus Schneider, William J. Fitzsimmons, Gaurav Khullar, Jessica Metti, Joel T. Dudley, Megan Nash, Nike Beaubier, Jianhui Wang, Chen Liu, Pei Hui, Anthony Muyombwe, Randy Downing, Jafar Razeq, Stephen M. Bart, Ardath Grills, Stephanie M. Morrison, Steven Murphy, Caleb Neal, Eva Laszlo, Hanna Rennert, Melissa Cushing, Lars Westblade, Priya Velu, Arryn Craney, Kathy A. Fauntleroy, David R. Peaper, Marie L. Landry, Peter W. Cook, Joseph R. Fauver, Christopher E. Mason, Adam S. Lauring, Kirsten St. George, Duncan R. MacCannell, Nathan D. Grubaugh

**Author notes:** These authors contributed equally (Tara Alpert, Anderson F. Brito, Erica Lasek-Nesselquist, Jessica Rothman, Andrew L. Valesano, Matthew J. MacKay). Senior author (Joseph R. Fauver, Christopher E. Mason, Adam S. Lauring, Kirsten St. George, Duncan R. MacCannell, Nathan D. Grubaugh).

## Abstract

The emergence and spread of SARS-CoV-2 lineage B.1.1.7, first detected in the United Kingdom, has become a global public health concern because of its increased transmissibility. Over 2500 COVID-19 cases associated with this variant have been detected in the US since December 2020, but the extent of establishment is relatively unknown. Using travel, genomic, and diagnostic data, we highlight the primary ports of entry for B.1.1.7 in the US and locations of possible underreporting of B.1.1.7 cases. Furthermore, we found evidence for many independent B.1.1.7 establishments starting in early December 2020, followed by interstate spread by the end of the month. Finally, we project that B.1.1.7 will be the dominant lineage in many states by mid to late March. Thus, genomic surveillance for B.1.1.7 and other variants urgently needs to be enhanced to better inform the public health response.

## Introduction

The rise of SARS-CoV-2 infections to unprecedented levels in the final months of 2020 has led to the evolution of several variants with concerning mutations or traits (Lauring and Hodcroft, 2021). One such variant designated B.1.1.7 (also 501Y.V1) was identified in the United Kingdom (UK) during the fall of 2020 and proceeded to predominate circulation in the region by early 2021 (Davies et al., 2021). Subsequent analyses suggested that B.1.1.7 quickly rose in frequency because it was approximately 43-90% more transmissible than other circulating lineages (Davies et al., 2021), resulting in a 0.4-0.7 increase in the effective reproductive rate in the UK (Volz et al., 2021). Further analysis of contact tracing data suggested that infections with the B.1.1.7 variant resulted in a 30-50% higher secondary attack rate (PHE, 2020). The B.1.1.7 SARS-CoV-2 variant is defined by 17 amino acid changes, including 8 changes in the spike protein (Rambaut et al., 2020). Of particular note is the N501Y mutation in the receptor-binding domain of the spike protein, which is predicted to increase binding to human angiotensin-converting enzyme 2 (ACE2) receptors (Starr et al., 2020) and is also a defining feature of other variants of concern such as B.1.351 and P.1 (Faria et al., 2021; Lauring and Hodcroft, 2021; Tegally et al., 2020). In addition, B.1.1.7 variants have a deletion in the spike gene (Δ69/70 HV) that may increase cell infectivity (Kemp et al., 2020) and has provided a serendipitous tracking method for B.1.1.7 by causing a “spike gene target failure” (SGTF) in the commonly used Thermo Fisher TaqPath COVID-19 Combo Kit (Borges et al., 2021; Volz et al., 2021). As of March 6, 2021, B.1.1.7 has been detected in 94 countries (cov-lineages.org/global_report.html), which raises concerns that B.1.1.7 will follow the trajectory that it took in the UK in other places around the world.

Current virus genomic surveillance across the United States (US) is uneven, creating uncertainty about the extent of international introductions, domestic spread, and community transmission of the SARS-CoV-2 variant. There have been 2672 reported COVID-19 cases associated with B.1.1.7 from 48 states (as of March 6, 2021)(CDC, 2021a), but these numbers are likely substantial underestimates. Approaches to enhance depth in certain populations and geographies may be required to address such uncertainty. Regardless, the existing framework limits the implementation of effective public health actions, such as targeted public health messaging and enhanced mitigation (Grubaugh et al., 2021), and allows B.1.1.7 to spread unimpeded (Galloway et al., 2021). The continued rise in B.1.1.7 cases may increase the burden on the US healthcare system and enable further evolution of mutations of public health concern (Wise, 2021).

Here, to investigate the locations of B.1.1.7 introductions into the US, identify significant surveillance gaps, and provide evidence for community transmission and domestic spread, we combined data from UK air travel into US airports, SARS-CoV-2 genomic sequencing, and clinical diagnostics. We identified where most B.1.1.7 introductions were most likely to have occurred and where surveillance for B.1.1.7 cases could be immediately supported. Combining our work with other analyses (Washington et al., 2021), we found that B.1.1.7 became independently established in parts of the US starting in early December 2020, and community transmission from some of these sources has already led to interstate spread. Finally, our TaqPath SGTF data indicates that the frequency of B.1.1.7 is rapidly rising, and we project that it will become the dominant SARS-CoV-2 lineage in many states by mid to late March, 2021. Thus, enhanced surveillance and control measures are urgently needed to mitigate B.1.1.7 leading to a resurgence of COVID-19 in the US.

## Results

### Locations of potential B.1.1.7 introductions from international travelers

During the early phases of the COVID-19 pandemic, international travel can seed the local establishment of new SARS-CoV-2 variants. Thus, to obtain a relative estimate of where B.1.1.7 introductions were most likely to occur, we analyzed air passenger travel volumes from December, 2020 coming into all US airports from the initial primary source of the variant - the UK (**Figure 1**; showing top 15 airports). This period, December, 2020, is when B.1.1.7 was rapidly expanding in the UK (Davies et al., 2021; Volz et al., 2021) and when we expect the first introductions to be occurring within the US. We found that the airports with the largest passenger volumes as a final destination from the UK were: New York JFK (7687), Los Angeles (3541), Newark Liberty (3371), Boston Logan (3216), Washington Dulles (2680), and Miami (2604) (**Figure 1C**, full list shown in **Data S1**). We then implemented a probabilistic choice behavior model to estimate where air passengers may travel after reaching their final airport destination (Huber et al., 2021). Thus by estimating the catchment of each airport, we created a county level risk map for where early international B.1.1.7 introductions may have occurred within the US (**Figure 1A-B**). After summing the risks across all counties, we predict the highest importation risks in New York, California, Florida, Texas, New Jersey, and Massachusetts.

**Figure 1.**
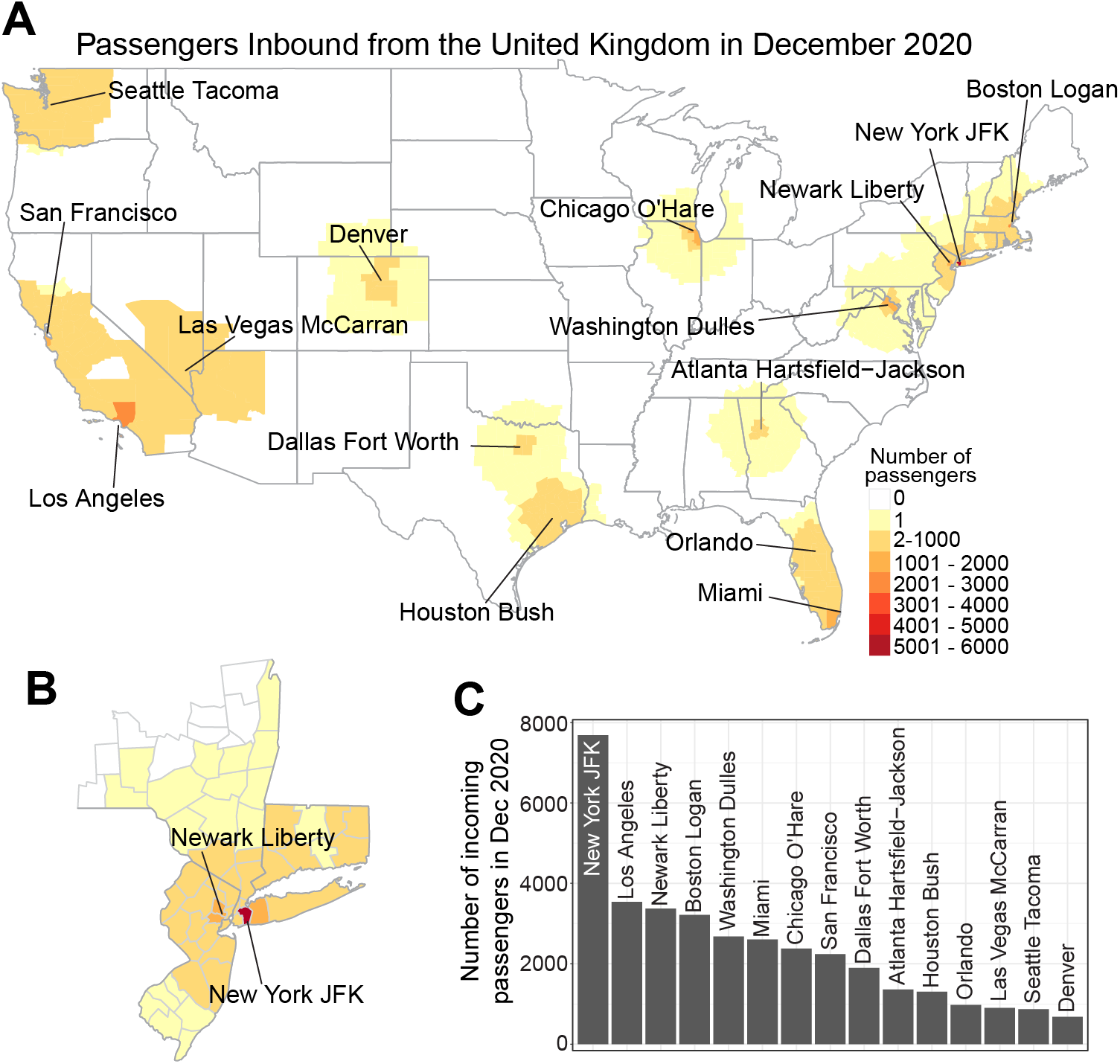
Identification of regions in the United States at risk for importation of B.1.1.7. **A**. County-level risk assessment of B.1.1.7 introductions from air passenger travelers entering US airports from the UK during December, 2020. Labeled are the top 15 airports in the US for passenger volumes from the UK (shown in **C**). The county-level heat map represents the probability of where passengers travel to after arriving at each airport (i.e. the airport catchment area, estimated using the Huff model multiplied by the total numbers of travelers entering each airport (see Methods). **B**. An expanded view of the counties in New York, New Jersey, and Connecticut to highlight the catchment of the large numbers of UK travelers entering the New York JFK and Newark Liberty airports. The same legend in **A** applies to **B. C**. The total number of passengers entering the top 15 US airports from the UK during December, 2020.

We focused our analysis on travelers entering the US from the UK. For completeness, however, we also obtained air passenger volumes from other countries that also reported B.1.1.7 cases in December, 2020: Germany, Denmark, and the United Arab Emirates (outbreak.info/situation-reports; **Table S1**). The total number of air passenger travelers from the UK to the US in December was 45,282, which is greater than the number coming from Germany (31,486), Denmark (5,544), and the United Arab Emirates (15,291); and the majority of the travelers arrived at similar airports (New York JFK, Newark Liberty, Chicago O’Hare, Washington Dulles, and Los Angeles). Finally, the frequency of B.1.1.7 was likely higher in the UK compared to other countries during this time, and thus the risk to travelers was greater. Therefore, we did not include other countries of origin in our flight-based importation risk assessment as they would unlikely change our findings of where B.1.1.7 importations would likely occur in the US (**Figure 1**).

### Genomic surveillance gaps and potential underreporting of B.1.1.7 cases

As of March 6, 2021, 2,672 B.1.1.7 cases from 48 states have been identified in the US (CDC, 2021a). These numbers, however, are likely significantly underreported as whole genome sequencing is needed for B.1.1.7 confirmation. To identify where B.1.1.7 cases may be disproportionately underreported during the early phases of B.1.1.7 emergence in the US (**Figure 2**), we evaluated the intensity of SARS-CoV-2 genomic surveillance for each state and compared that to our flight-based risk estimates of where early B.1.1.7 outbreaks may have occurred (**Figure 1**).

**Figure 2.**
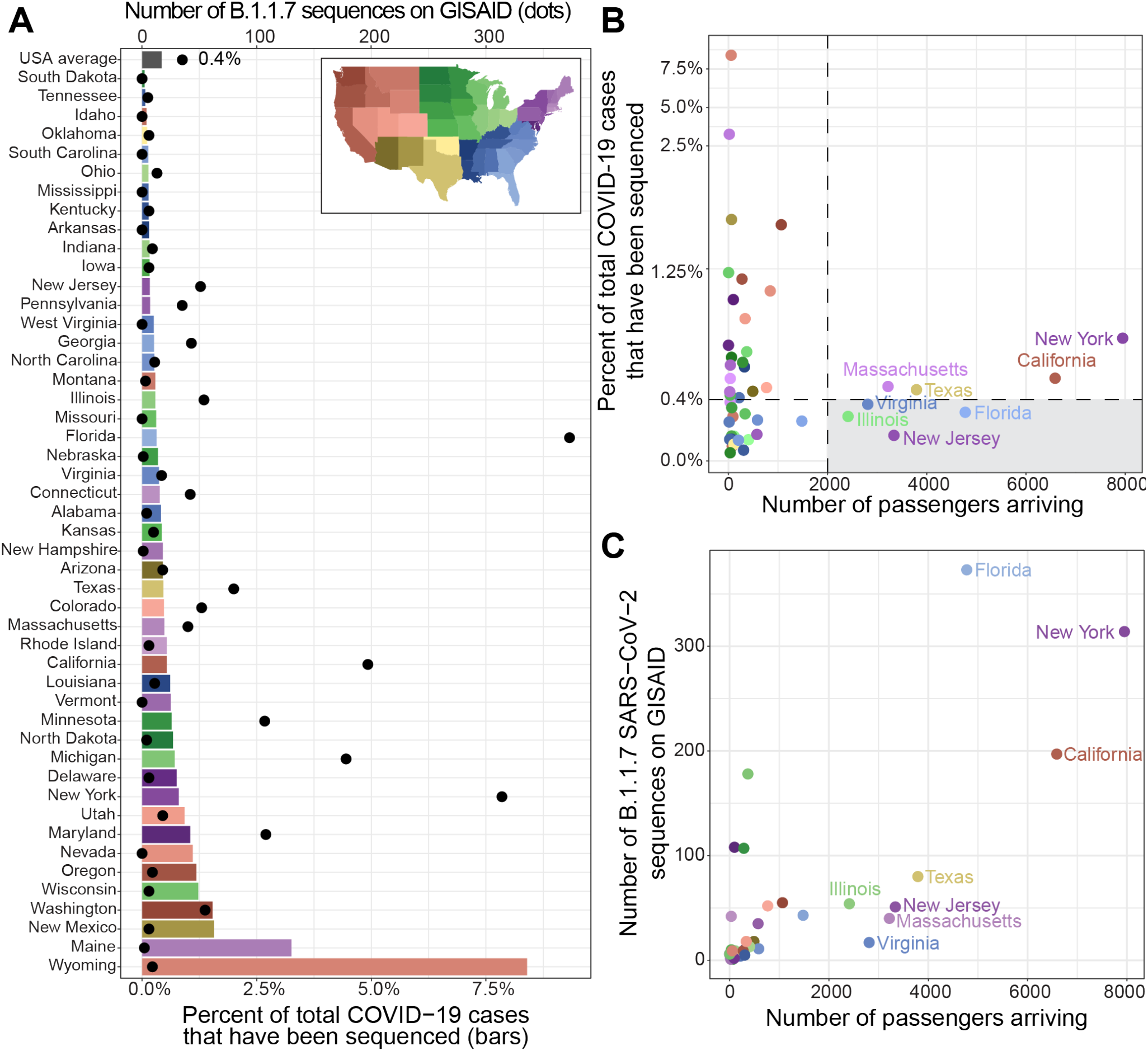
Identification of genomic surveillance gaps and regions that may be disproportionately underreporting B.1.1.7. **A**. Bar plot represents the percentage of cases in each state from Dec 2020 to Feb 2021 (bottom x-axis; sourced from covidtracking.com/data) that have sequences uploaded to GISAID.org (accessed on Mar 4, 2021). Bars are colored according to region (legend, top right). The number of B.1.1.7 sequences for each state (top x-axis; black dots) was determined by the Pangolin lineage assignment in the GSIAID.org metadata. **B**. Total number of passengers arriving from the UK in Dec 2020 to each state in the continental US (data from Huff model in Figure 1) is plotted against the percent of sequenced COVID-19 cases. The horizontal dashed line represents the US average (0.43%) for sequenced cases. States sequencing below the US average with more than 2,000 passengers (vertical dashed line) are at risk for underreporting B.1.1.7 (grey box). **C**. Number of B.1.1.7 SARS-CoV-2 sequences available on GISAID.org for each state. Points are colored according to region (legend from panel **A**). The data used to create this figure are listed in **Data S1**.

We started by evaluating the percent of sequenced SARS-CoV-2 clinical samples relative to the number of reported COVID-19 cases (**Figure 2A, Figure S1**). For this, we downloaded (***1***) all SARS-CoV-2 genomes available on GISAID (gisaid.org; accessed on March 4, 2021) with “USA” listed as a location and (***2***) the total number of new COVID-19 cases for each state from December 2020 to February 2021 (covidtracking.com; accessed on March 4, 2021; **Data S1**). For this three month period, which was crucial for B.1.1.7 introductions and establishment, we found that only 0.43% of the US COVID-19 cases sequenced and posted on GISAID. As sample testing, transport, sequencing, analysis, and data submission can take multiple days to weeks, more data from this time period will likely be available in the near future. This delay can be seen for most states, which have a relatively lower percentage of sequenced cases available for February than January (**Figure S1**). Still, there are 24 states with less than 0.4% of the COVID-19 cases with available SARS-CoV-2 sequences during December 2020 to February 2021, including 9 states that have not submitted any B.1.1.7 sequences (**Figure 2A, Figure S1, Data S1**).

While a low fraction of sequenced COVID-19 cases will certainly hinder the detection of B.1.1.7 and other variants of concern, these data alone may not indicate where B.1.1.7 may be disproportionately underreported. Therefore we compared our risk estimates of B.1.1.7 introductions using air passenger volumes from the UK to all SARS-CoV-2 (**Figure 2B, Data S1**) and B.1.1.7 (**Figure 2C, Data S1**) genomes sequenced per state. Of the states receiving more than 2,000 air passengers from the UK, we found that COVID-19 cases from New Jersey (0.17% of cases sequenced), Illinois (0.29%), Florida (0.32%), and Virginia (37%) have been sequenced below the 0.43% national average (**Figure 2B**). In Florida, many of the available SARS-CoV-2 sequences were targeted using TaqPath SGTF results, and thus they have sequenced 373 B.1.1.7 genomes to date, the most in the country (**Figure 2C**). In places like New Jersey, Illinois, and Virginia, however, if SARS-CoV-2 genomic surveillance could be increased it might determine if B.1.1.7 cases are disproportionately underreported as compared to New York and California.

### Phylogenetic evidence for multiple B.1.1.7 introductions and interstate spread

Our travel data indicated that we should have observed many separate and sustained B.1.1.7 introductions within the US from the UK and perhaps other international locations where the variant may be circulating. To investigate if some of these introductions led to community transmission and/or interstate spread within the US, we combined our SARS-CoV-2 sequencing data from the US Centers for Disease Control and Prevention (CDC, 568 sequences), Yale University (116), University of Michigan (45), and New York State Department of Health (41). Our phylogenetic analysis of these sequences suggests that there were many separate introductions into the US that led to secondary transmission, and that some sustained introductions within states were likely from domestic spread (**Figure 3**).

**Figure 3.**
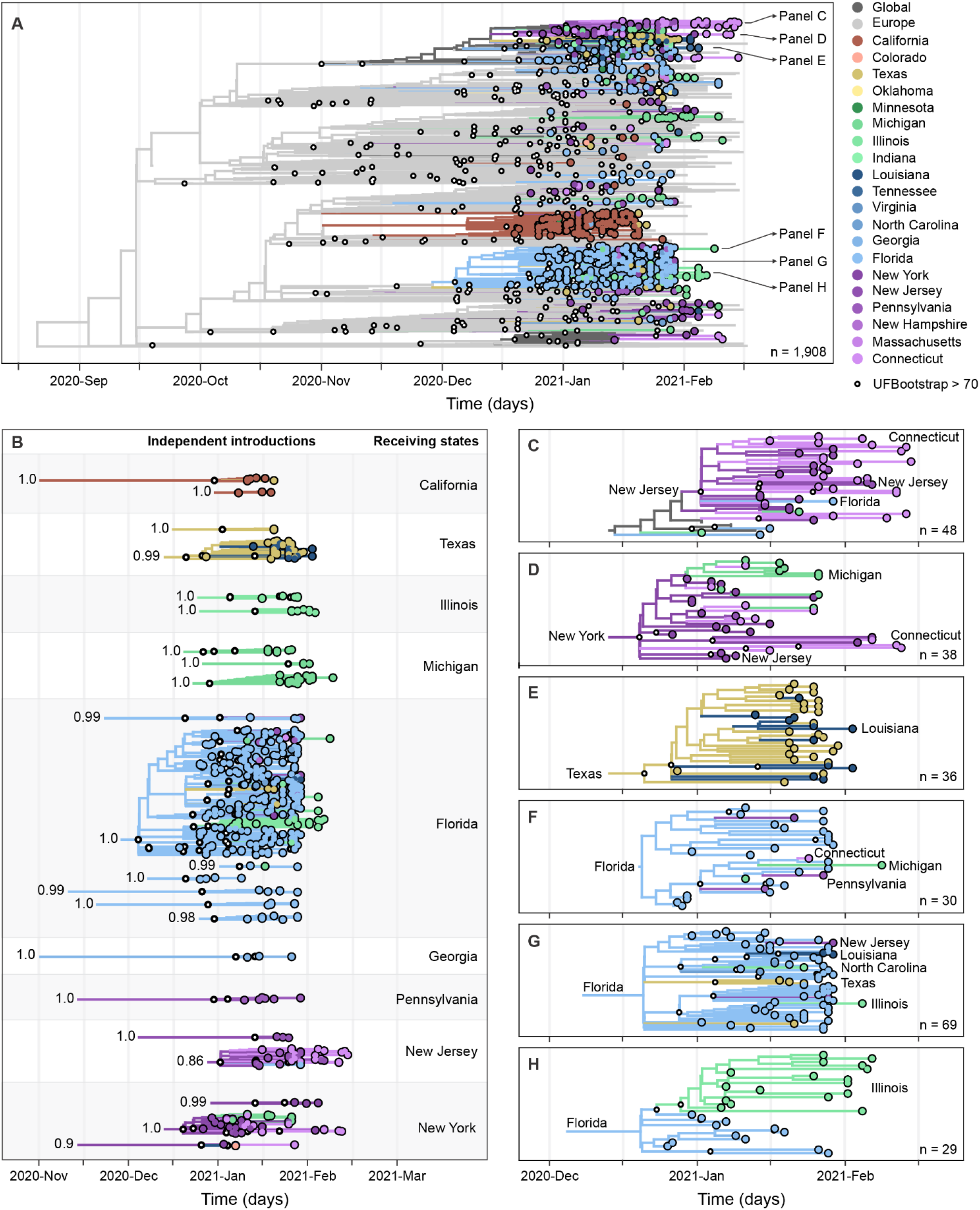
Multiple introductions, domestic spread, and community transmission of B.1.1.7 SARS-CoV-2 in the United States. **A**. Maximum likelihood phylogeny of B.1.1.7, including 1,908 representative genomes from the US, Europe, other global locations. Tree topology and bootstrap values obtained using IQ-Tree 1.6.12, with time scale inferred by TreeTime 0.8.0, discrete state reconstruction inferred using BEAST v1.10, and data integration, and visualization using baltic 0.1.5. The tree was rooted using a P.1 genome (Brazil/AM-20842882CA/2020) as an outgroup (not shown in this plot). **B**. Exploded tree layout, highlighting clades with 3 or more taxa, UFBoot > 70 (small circles), and US ancestral state probability at MRCA > 0.7 (values at the root), representing independent international introductions of B.1.1.7 into distinct regions of the US, based on the same phylogenetic tree shown in (**A**). A list of international transitions to the US can be found in **Data S1. C-H**. Time-informed maximum likelihood phylogeny of distinct B.1.1.7 clades showing instances of intra-region (**C, D, E, G**) and inter-region (**D, H**) domestic spread. The list of SARS-CoV-2 sequences used in this study and author acknowledgements can be found in **Data S2**. Supporting phylogenetic analysis can be found in **Figures S2-5**. For comparison, an interactive phylogenetic tree, inferred using IQ-Tree and TreeTime only, can be accessed from our custom Nextstrain build: nextstrain.org/community/grubaughlab/CT-SARS-CoV-2/paper5.

We generated or received permission to use 770 B.1.1.7 genomes collected from December 19, 2020 to February 14, 2021 from the following states: Florida (267), California (133), Illinois (64), New York (49), Michigan (48), Connecticut (47), Texas (45), New Jersey (35), Georgia (33), Pennsylvania (11), North Carolina (10), Louisiana (9), Indiana (4), Massachusetts (4), Minnesota (3), Tennessee (3), Oklahoma (2), Colorado (1), New Hampshire (1), and Virgina (1). From a larger dataset of 101,079 B.1.1.7 genomes available up to February 26, 2021, we generated a smaller set of 8,864 B.1.1.7 genomes from 59 countries (7,589 from international locations and 1,275 from the US, **Figure S2**) using a subsampling method based on COVID-19 incidence (see Methods). We further reduced this tree to a representative set of 1,908 genomes, which included our data and 1,139 B.1.1.7 genomes available from the UK and other countries to infer a time-scaled maximum-likelihood phylogenetic tree. We subsequently conducted discrete phylogeographic analysis of our fixed time-scaled maximum-likelihood tree using a Bayesian approach (Dellicour et al., 2020; Lemey et al., 2009) to identify descending clades that likely represent distinct introductions into US states (**Figure 3**). Furthermore, to identify B.1.1.7 introductions that likely led secondary community transmission, we limited our analysis to descending clades originating in the US with (***1***) 3 or more sequences, (***2***) > 70 bootstrap support, and (***3***) > 70% discrete state probability within the US at the most recent common ancestor (MRCA). Based on these criteria and by performing ancestral trait reconstruction using BEAST, we found 23 distinct B.1.1.7 introductions in the US that led to secondary transmission (**Figure 3B**). From these descending clades, we estimate that the median times to the MRCA (tMRCA), and, by proxy, the estimated times in which B.1.1.7 became locally established, occurred throughout early December to January (**Figure 3B, Data S1**). Specifically, from clades with more than 15 sequences, we estimate B.1.1.7 establishment in Florida by early December (clade size = 243; 90% CI = 2020-11-25 to 2020-12-11), New York by mid December (38; 2020-12-16, 2020-12-23), Texas mid December (36; 2020-12-16 to 2020-12-26), Michigan by late December (18; 2020-12-24 to 2020-12-30), and New Jersey/Connecticut by early January (34; 2020-12-28 to 2021-01-06; **Data S1**). By comparison, Washington et al. estimated the tMRCA for the Florida clade to be December 3^rd^ (95% highest posterior probability: 2020-11-22 to 2020-12-11) (Washington et al., 2021), which is within days of our estimate. They also document a sustained B.1.1.7 introduction into California during early December (Washington et al., 2021), a clade with 91 genomes that can be visualized in **Figure 3A**, which did not get included as an introduction in our analysis as it did not meet our criteria of > 70 bootstrap support.

We also discovered several instances of likely interstate B.1.1.7 spread that occured between December, 2020, and January, 2021 (**Figure 3C-H**). For example, we found regional spread within the New York, New Jersey, and Connecticut (**Figure 3C-D**), which is expected based on their connectedness among travelers (**Figure 1B**) and that New York was a regional “hub” for SARS-CoV-2 spread during the early pandemic (Gonzalez-Reiche et al., 2020; Maurano et al., 2020). We also found evidence for regional spread between Texas and Louisiana (**Figure 3E**) and from Florida to several other states in the Southeast US (**Figure 3G**), which is further supported by independent findings of a “Southeast” B.1.1.7 clade (Washington et al., 2021). Finally, our data suggests that out of region spread has also occurred from the New York/New Jersey/Connecticut region to Michigan (**Figure 3D**; tMRCA 90% CI: 2020-12-24 to 2021-01-07) and from Florida to Illinois (**Figure 3H**; 2020-12-25 to 2021-01-01). Additional examples can be found by exploring our full dataset (nextstrain.org/community/grubaughlab/CT-SARS-CoV-2/paper5).

Our estimates of international and domestic introductions, however, can be significantly influenced by sampling biases and gaps, sequencing and processing errors, and the imperfection of estimating transitions between locations among sequences with low genetic diversity. Moreover, based on the low rate of sequencing of COVID-19 cases in the US (**Figure 2A**), the true number of international and domestic introductions are likely significantly higher than what we reported. Importantly, though, our results inform us that many sustained B.1.1.7 introductions occurred throughout the country, with some likely weeks before the first COVID-19 cases associated with B.1.1.7 were reported in the US during late December (Zimmer and Pietsch, 2020).

### Diagnostic evidence for increased community transmission of B.1.1.7

Our phylogenetic analysis and those by our colleagues (Washington et al., 2021) indicate that the B.1.1.7 introductions that led to community transmission in the US began around early December 2020 (**Figure 3**). To investigate if B.1.1.7 has increased in frequency since these introductions, we used TaqPath assay SGTF results from COVID-19 clinical samples (**Figure 4**). As the spike gene Δ69/70 HV deletion in B.1.1.7 genomes causes SGTF results when using the TaqPath assay, tracking the occurrence of SGTF can provide an indirect measure of changes in B.1.1.7 population frequency (Borges et al., 2021; Volz et al., 2021). Our SGTF data suggest that B.1.1.7 is increasing in frequency in all four states investigated (Connecticut, New York, New Jersey, and Illinois) and will likely become the majority SARS-CoV-2 lineage between mid to late March, 2021, in line with other estimates (Washington et al., 2021).

**Figure 4.**
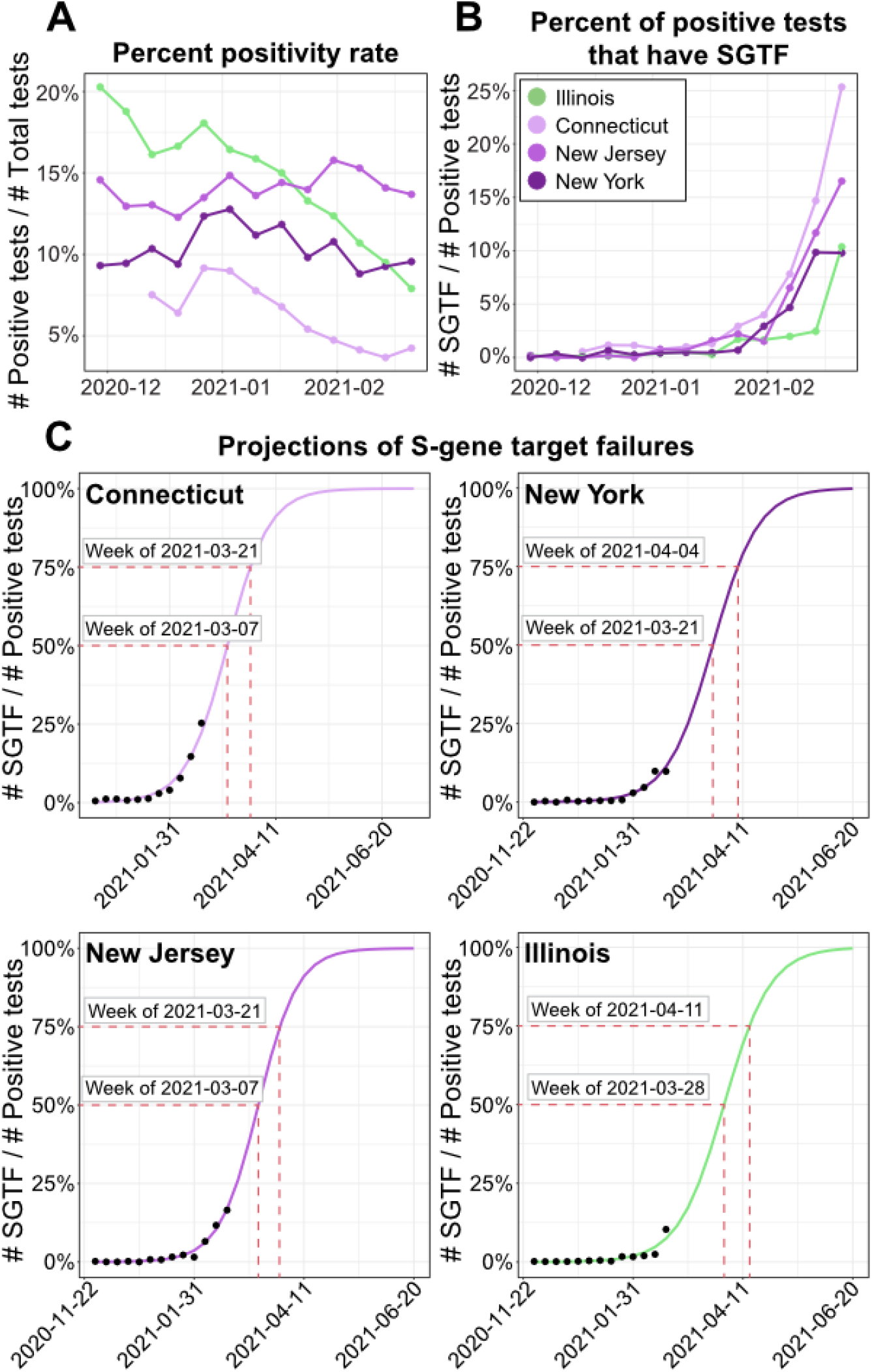
Increasing frequency of weekly spike gene target failure (SGTF) results across four US states. **A**. The weekly positivity rate of SARS-CoV-2 testing for four states (legend, panel **B**) since the first week of December, 2020, calculated as the number of positive tests (including SGTF) divided by total tests. **B**. The percentage of weekly positive tests which have SGTF are shown for the same time-period and states from **A (legend, top left). C**. The weekly percent SGTF data from panel B fit to a logistic regression model (see Methods) to project the week in which we estimate SGTF results, and by proxy B.1.1.7, will cross the 50% and 75% thresholds for each state population. The color schemes shown in panels A-C match the color schemes used in Figures 2-3. The data used to create this figure are listed in **Data S1**.

We obtained 422,330 TaqPath COVID-19 RT-PCR test results performed by Tempus and Yale New Haven Hospital clinical diagnostic laboratories from December 2020 through February 2021 on nasal swab samples collected from 4 states: Illinois (183,077 tests), Connecticut (139,403), New Jersey (58,675), and New York (41,175). The weekly SARS-CoV-2 test positivity rate varied across the states and months, but all were high (>8%) in late December to early January (**Figure 4A**; **Data S1**). From our tested samples from Illinois and Connecticut, we saw a notable decrease in the percent of SARS-CoV-2 positive tests which follow the national trends.

Among the positive tests, the frequency of SGTF results remained low (<2%) until late January when they dramatically rose to 10-25% across all states by late February (**Figure 4B**). During January, 2021, only 47% (54/116) of the SGTF samples presented in **Figure 4B** that we sequenced were identified as B.1.1.7, while the majority were SARS-CoV-2 lineage B.1.375, which also has the spike gene Δ69/70 HV deletion that causes SGTF (Larsen and Worobey, 2020; Moreno et al., 2021). In February, however, we found that 90% (254/282) of the SGTF samples that we sequenced were B.1.1.7, including 100% (147/147) from February 15-23. Thus, the proportion of SARS-CoV-2 positive samples generating a SGTF result are on the rise across several states, and SGTF is becoming a near-direct measure of B.1.1.7.

Next we wanted to estimate when we should expect B.1.1.7 to become the majority SARS-CoV-2 lineage among the tested populations represented by our data. We fit our SGTF data to a logistic regression model showing an exponential increase in the percent of SGTF results among the positive tests, representing an exponential growth of B.1.1.7 across all sites (**Figure 4C**). From this analysis, we estimate that SGTF results will reach the 50% threshold during the week of March 7, 2021, in Connecticut, March 21 in New York, and March 28 in Illinois and New Jersey. Furthermore, we estimate that SGTF results, and by proxy B.1.1.7, will reach the 75% threshold of positive cases about 2 weeks later. Thus, by mid-April, parts of the COVID-19 pandemic in the US will be dominated by the transmission properties of B.1.1.7.

## Discussion

Despite travel restrictions and increased testing requirements, we found evidence for a large number of independent B.1.1.7 introductions into the US, many of which have led to secondary community transmission. Incoming air passenger volumes from the UK predict that New York, California, and Florida would be at highest risk for importation, and high numbers of B.1.1.7 sequences from these states agree with that hypothesis. Indeed, our phylogenetic analyses suggest that in addition to separate introduction events, B.1.1.7 became independently established across the country starting in early December, 2020, weeks before the first reported case in the US on December 29, 2020 (Zimmer and Pietsch, 2020). Around the same time, we found several examples of within-region interstate spread of B.1.1.7 in the northeast, southeast, and southern US, and some examples of out-of-region spread from the northeast and southeast to the midwest. This period of “silent” spread across the US is reminiscent of the early 2020 COVID-19 pandemic in the US when diagnostic testing was low (Fauver et al., 2020). Overall, our data highlight the relative ease with which SARS-CoV-2 variants can spread undetected throughout the US, particularly in areas where genomic surveillance efforts are minimal.

COVID-19 cases associated with the B.1.1.7 variant are likely significantly underreported across the US. This is because, as a whole, only about 0.43% of the COVID-19 cases in the US were sequenced from December 2020 to February 2021, which included the period with the highest case rates in the country. The sequencing capacity is highly variable across the country, and our travel data helps to identify regions which may be disproportionately underreporting cases of B.1.1.7 and where it would be prudent to immediately prioritize variant surveillance. States such as New Jersey, Illinois, and Virginia received moderately high levels of UK travel, yet reported a low proportion of sequenced COVID-19 cases from recent months, presenting the likelihood that B.1.1.7 is significantly underdetected in these regions.

It is estimated that 5% of the COVID-19 cases should be sequenced to detect emerging variants when they exist at a prevalence of 0.1% to 1.0% (Vavrek et al., 2021). With the nation-wide decrease in COVID-19 cases since reaching a new peak in early January 2021 and the initiatives to increase the capacity of SARS-CoV-2 sequencing within state public health laboratories (CDC, 2021b), there should be a considerable increase in the proportion of COVID-19 cases sequenced in the US during coming months. However, we should not expect an immediate jump to that 5% threshold, and we will need to supplement sequencing with other more conventional laboratory approaches. In the UK, the frequency of B.1.1.7 was primarily tracked across locations using SGTF results from the TaqPath COVID-19 PCR assay (Borges et al., 2021; Volz et al., 2021). Here we also demonstrate how SGTF data can help to provide insights on B.1.1.7 population frequency changes, and how these results can help to prioritize samples for sequencing. In fact, Florida also received a high volume of UK travel (ranked #3 in the US) and has a low overall rate of virus sequencing (0.32% of cases sequenced, ranked #29), but targeted sequencing of SGTF samples has helped identify a large number of B.1.1.7 cases in this state (373, ranked #1)(Washington et al., 2021). While TaqPath SGTF results are not definitive for B.1.1.7 (Larsen and Worobey, 2020; Moreno et al., 2021), as its frequency climbs, SGTF data becomes closer to a proxy for B.1.1.7 presence. Thus, the TaqPath clinical diagnostic assay, plus research-use only PCR assays that are more specific for B.1.1.7 detection (and that can also detect other current variants of concern) (Vogels et al., 2021), could provide immediate data to guide public health decision-making, especially in areas where B.1.1.7 cases may be disproportionately underestimated.

Our study has important limitations. First, our importation risk analysis did not account for the likelihood of transmission among the different regions in the US. As COVID-19 cases were at or near their peak across the country, our assumption was that transmission potential was high everywhere, and that the numbers of potentially infected travelers was a more significant factor. In reality, local conditions and behaviors play an important role for B.1.1.7 establishment, and could explain why B.1.1.7 cases are low in some states as opposed to surveillance deficiencies. Second, while we provide substantial evidence for several independent introductions, increased community transmission, and domestic spread, the significant undersampling that we discuss throughout this manuscript highlights that these events are likely underestimated. As we generate more data, we will be able to reveal additional insights into the patterns of B.1.1.7 spread across the US and the level to which B.1.1.7 will achieve dominance in different regions.

The SARS-CoV-2 B.1.1.7 variant of concern has become established in many states within the US. Our data and those of our colleagues (Washington et al., 2021) indicate that B.1.1.7 is expanding at an exponential rate, and that it will be the dominant SARS-CoV-2 lineage in many places across the US by March or April, 2021. While surveillance gaps across the US mean that some communities do not have direct evidence for local B.1.1.7 emergence, it should be assumed that community B.1.1.7 transmission is widespread. With several US states announcing loosening restrictions on gatherings (including restaurants), restrictions on travel, and/or mask requirements, and with sufficient COVID-19 vaccine coverage to reach population immunity still many months away (summer/fall, 2021), the near-term impact of B.1.1.7 may be significant. We must use this opportunity to reinforce communications and messaging surrounding the importance of mitigation measures to prevent this variant from exacerbating an already crippling pandemic (Grubaugh et al., 2021). In reality, it is difficult to implement new measures without supporting data, especially if they impact schools or businesses. Thus, increasing surveillance for B.1.1.7 and other variants through sequencing and more conventional methods should be made a high priority (CDC, 2021b).

### Experimental Model and Subject Details

#### Ethics Statement

The Institutional Review Board from the Yale University Human Research Protection Program determined that the RT-qPCR testing and sequencing of de-identified remnant COVID-19 clinical samples obtained from clinical partners conducted in this study is not research involving human subjects (IRB Protocol ID: 2000028599).

Residual nasopharyngeal and saliva specimens from individuals who tested positive for SARS-CoV-2 by RT-PCR were obtained from the Michigan Medicine Clinical Microbiology Laboratory, University (of Michigan) Health Services, and LynxDx (Ann Arbor, MI). This work was approved by the University of Michigan Institutional Review Board (IRB Protocol ID: HUM185966), Expanded sequencing in January 2020 was performed as part of a public health investigation.

Residual portions of respiratory specimens from individuals who tested positive for SARS-CoV-2 by RT-PCR were obtained from the Wadsworth Center and partnering clinical laboratories. This work was approved by the New York State Department of Health Institutional Review Board, under study numbers 02-054 and 07-022.

This activity was reviewed by CDC and was conducted consistent with applicable federal law and CDC policy.

## Methods

### Flight volumes and maps

The flight travel volume data were provided by OAG Aviation Worldwide Ltd. OAG Traffic Analyser, Version 2.5.11 2020 (analytics.oag.com/analyser-client/home; accessed 2020-02-22). Travel volume numbers are modeled estimates based on ticket sales and reporting from airline carriers. Travel volume represents the aggregate number of passenger journeys, not necessarily unique individuals. A subset of the available data was used only to capture flights whose origin was the UK and whose final destination was an airport in the US for flights that occurred in December 2020.The map presented in **Figures 1A-B**, which shows the approximate final destinations of the estimated number of people per county who flew into the top 15 airports in the US on flights inbound from the UK in December 2020 and overlaid onto a map of the US, was generated using R, with the maps, choroplethr, and ggplot2 packages (Becker et al., 2018; Wickham, 2016). The UK inbound flight volume data per airport in the US are displayed in Figure 1C. These data were also used to calculate the total travelers from the UK per state and was used to generate **Figures 2B-C**.

### Sample selection, screening, and SARS-CoV-2 sequencing

#### Yale University

##### Sample selection and RNA extraction

Samples were received in partnership with various clinical laboratories as either purified RNA or original nasal swab in viral transport media. Samples were screened for S-gene target failure (SGFT) using the Thermo Fisher TaqPath COVID-19 Combo Kit diagnostic assay prior to receipt at Yale. Nucleic acid was extracted from original samples (300 μL) using the MagMAX viral/pathogen nucleic acid isolation kit (Thermo Fisher) and eluted into 75 μL. All RNA was then screened again using an assay developed by our laboratory that is specific for variants of concern (Vogels et al., 2021). Samples identified by the screen as potential variants were then prioritized for sequencing. Multiple extraction controls were included for each RNA extraction batch and tested negative for SARS-CoV-2 RNA by the same assay.

##### Oxford Nanopore library preparation and sequencing

RNA extracted from positive samples served as the input for an amplicon-based approach for sequencing on the Oxford Nanopore Technologies (ONT; Oxford, United Kingdom) MinION (Quick et al., 2017). Sequencing libraries were prepared using the ONT Ligation Sequencing Kit (SQK-LSK109) and the ONT Native Barcoding Expansion pack as described in the ARTIC Network’s protocol with V3 primers (IDT) (Quick, 2020) with the following modifications: cDNA was generated with SuperScriptIV VILO Master Mix (Thermo Fisher Scientific, Waltham, MA, USA), all amplicons were generated using 35 cycles of amplification, amplicons were then normalized to 15 ng for each sample, end repair incubation time was increased to 25 min followed by an additional bead-based clean up, and all clean up steps used a ratio of 1:1 beads:sample. No-template controls were introduced for each run at the cDNA synthesis and amplicon synthesis steps and were taken through the entire library preparation and sequencing protocol to detect any cross-contamination. For each control in each run, less than 1,000 total reads were observed. A subset of reads in control samples aligned to the SARS-CoV-2 genome, although no position of the genome had greater than 20 reads i.e. enough data to influence the generation of a consensus genome. 25 ng of the final library was loaded on a MinION R9.4.1 flow cell and sequenced for approximately 8-10 hours.

##### Bioinformatics processing

The RAMPART application from the ARTIC Network was used to monitor approximate genome coverage for each sample and control in real time during the sequencing run (github.com/artic-network/rampart). Fast5 files were basecalled using the Guppy basecaller 4.4.0 fast model and consensus genomes were generated according to the ARTIC bioinformatic pipeline (artic.network/ncov-2019/ncov2019-bioinformatics-sop.html) which uses Nanopolish to call variants (Loman et al., 2015). A threshold of 20x coverage was required for each amplicon to be included in the consensus genome.

#### University of Michigan

##### Sample selection, RNA extraction, reverse transcription, and genome amplification

All available specimens in the month of January were prepared for sequencing. Residual transport media or saliva was centrifuged at 1200 x g. and aliquoted. For nasopharyngeal and sputum specimens, RNA was extracted with the Invitrogen PureLink Pro 96 Viral RNA/DNA Purification Kit (200 μL of input sample eluted in 100 µL) or the QIAamp Viral RNA Mini kit (140 µL of input sample eluted in 50 µL). For saliva specimens, RNA was extracted with the Thermo Fisher MagMAX Viral RNA Isolation Kit (200 µL of input sample eluted in 50 µL). Extracted RNA was reverse transcribed with SuperScript IV (Thermo Fisher). For each sample, 1 µL of random hexamers and 1 µL of 10 mM dNTP were added to 11 µL of RNA, heated at 65 deg°C for 5 min, and placed on ice for 1 min. Then a reverse transcription master mix was added (4 µL of SuperScript IV buffer, 1 µL of 0.1M DTT, 1 µL of RNaseOUT RNase inhibitor, and 1 µL of SSIV reverse transcriptase) and incubated at 42 °C for 50 min, 70 °C for 10 min, and held at 4 °C. SARS-CoV-2 cDNA was amplified in two multiplex PCR reactions with the ARTIC Network version 3 primer pools and protocol. Viral cDNA was amplified with the Q5 Hot Start High-Fidelity DNA Polymerase (NEB) with the following thermocycler protocol: 98 °C for 30 s, then 35 cycles of 98 °C for 15 s, 63 °C for 5 min, and final hold at 4 °C. Reaction products for a given sample were pooled together in equal volumes.

##### Illumina library preparation and sequencing

Pooled PCR product was purified with 1X volume of AMPure beads (Beckman-Coulter). Sequencing libraries were prepared with the NEBNext Ultra II DNA Library Prep Kit (NEB) according to the manufacturer’s protocol. Barcoded libraries were pooled in equal volume and extracted with a 1% agarose gel to remove adapter dimers. Pooled libraries were quantified with the Qubit 1X dsDNA HS Assay Kit (Thermo Fisher). Libraries were sequenced on an Illumina MiSeq (v2 chemistry, 2×250 cycles) at the University of Michigan Microbiome Core facility. Reads were aligned to the Wuhan-Hu-1 reference genome (GenBank MN908947.3) with BWA-MEM version 0.7.15. Sequencing adaptors and amplification primer sequences were trimmed with iVar 1.2.1. Consensus sequences were called with iVar 1.2.1 by simple majority at each position (>50% frequency), placing an ambiguous N at positions with fewer than 10 reads.

##### Oxford Nanopore library preparation and sequencing

After multiplex PCR amplification, libraries were prepared for sequencing with the Oxford Nanopore Technologies MinION using the ARTIC Network version 3 protocol (Quick, 2020). Samples were prepared in batches of 24 with one-pot native barcoding. Pooled PCR products were diluted in nuclease-free water with a dilution factor of 10. Amplicon ends were prepared for ligation with the NEBNext Ultra II End Repair/dA-Tailing Module (NEB). Unique barcodes (Oxford Nanopore Native Barcoding Expansion kits) were ligated per sample with the NEB Blunt/TA Ligase Master Mix. After barcoding, reactions were pooled together in equal volumes and purified barcoded amplicons with 0.4X volume of AMPure beads. Oxford Nanopore sequencing adapters were ligated with the NEBNext Quick Ligation Module (NEB) and the library was purified with 1X volume of AMPure beads. Final libraries were quantified with the Qubit 1X dsDNA HS Assay Kit (Thermo Fisher). Each library (15-20 ng) was loaded onto a flow cell (FLO-MIN106) and sequenced with the MinION.

##### Bioinformatics processing

Sequencing progress was monitored with RAMPART. Basecalling was performed with Guppy v4.0.14 and consensus genomes were called using the ARTIC Network bioinformatics pipeline (artic.network/ncov-2019/ncov2019-bioinformatics-sop.html).

#### New York State Department of Health, Wadsworth Center

##### Sample selection and RNA extraction

Respiratory swabs in viral transport medium previously identified as SARS-CoV-2 positive by real-time RT-PCR were selected for sequencing, and included specimens received and tested in the Wadsworth Virology Laboratory and those submitted by clinical laboratories. An enhanced surveillance program was initiated in December 2020 and included retrospective sequencing of positive samples dating back to September 2020. Samples were generally required to have real-time Ct values less than 30 and minimal residual volumes of 100 µL. Most nucleic acid extractions were performed on a Roche MagNAPure 96 with the Viral NA Small Volume Kit (Roche, Indianapolis, IN) with 100µL sample input and 100µL eluate. Samples of special concern with Ct values in the low 30s were extracted on a NUCLISENS easyMAG instrument (bioMerieux, Durham, NC) with 1,000 µL sample input and 25 µL eluate.

##### Illumina library preparation and sequencing

Extracted RNA was processed for whole genome sequencing with a modified ARTIC protocol (artic.network/ncov-2019) in the Applied Genomics Technology Core at the Wadsworth Center. Briefly, cDNA was synthesized with SuperScript™ IV reverse transcriptase (Invitrogen, Carlsbad, CA, USA) and random hexamers. Amplicons were generated by pooled PCR with two premixed ARTIC V3 primer pools (Integrated DNA Technologies, Coralville, IA, USA). Additional primers to supplement those showing poor amplification efficiency (github.com/artic-network/artic-ncov2019/tree/master/primer_schemes/nCoV-2019) were added separately to the pooled stocks. PCR conditions were 98°C for 30 seconds, 24 cycles of 98°C for 15 seconds/63°C for 5 minutes, and a final 65°C extension for 5 minutes. Amplicons from pool 1 and pool 2 reactions were combined and purified by AMPure XP beads (Beckman Coulter, Brea, CA, USA) with a 1X bead-to-sample ratio and eluted in 10mM Tris-HCl (pH 8.0). The amplicons were quantified using Quant-IT™ dsDNA Assay Kit on an ARVO™ X3 Multimode Plate Reader (Perkin Elmer, Waltham, MA, USA). Illumina sequencing libraries were generated using the Nextera DNA Flex Library Prep Kit with Illumina Index Adaptors and sequenced on a MiSeq instrument (Illumina, San Diego, CA, USA).

##### Oxford Nanopore library preparation and sequencing

RNA was processed using the same ARTIC V3 protocol as described for Illumina library preparation. MinION libraries for up to 24 samples were generated according to the COVID-19 PCR tiling protocol (ONT). Native barcodes (ONT EXP-NBD104 and EXP-NBD114) were ligated to each DNA sample with NEB Blunt/TA Ligase Master Mix. Amplicons were pooled and purified using 0.4X AMPure XP beads and short-fragment buffer (ONT EXP-SFB001). Oxford Nanopore sequencing adapters were ligated with NEBNext Quick Ligation Module (NEB) and libraries were purified with 0.4X AMPure XP beads. About 15-25ng of each library was loaded on a FLO-MIN106 flowcell and sequenced with the MinION. Basecalling was performed by Guppy v4.2.3.

##### Bioinformatics processing

Illumina libraries were processed with ARTIC nextflow pipeline (github.com/connor-lab/ncov2019-artic-nf/tree/illumina, last updated April 2020). Briefly, reads were trimmed with TrimGalore (github.com/FelixKrueger/TrimGalore) and aligned to the reference assembly MN908947.3 (strain Wuhan-Hu-1) by BWA (Li & Durbin 2010). Primers were trimmed with iVar (Grubaugh et al. 2018) and variants were called with samtools mpileup function (Li et al. 2009), the output of which was used by iVar to generate consensus sequences. Positions were required to be covered by a minimum depth of 50 reads and variants were required to be present at a frequency ≥0.75. Consensus sequences were generated by the ARTIC bioinformatic pipeline v1.1.3 with Medaka variant calling (artic.network/ncov-2019/ncov2019-bioinformatics-sop.html) for Oxford Nanopore libraries.

#### Centers for Disease Control and Prevention

COVID-19 clinical samples sequenced by the CDC or sponsored by the CDC were generated for other purposes and shared with our teams via GISAID (gisaid.org). See elsewhere for some of the sequencing methodology and data processing (Washington et al., 2021).

### Clinical TaqPath COVID-19 RT-PCR testing and SGTF determination

Routine clinical COVID-19 diagnostic testing was performed in Clinical Laboratory Improvement Amendments of 1988 (CLIA) regulated laboratories at Yale University, Yale New Haven Hospital, and Tempus Labs following Emergency Use Authorization protocols submitted to the US Food and Drug Administration for use of the Applied Biosystems TaqPath COVID-19 Combo Kit (catalog number A47814). SGTF results were defined as any SARS-CoV-2 positive sample with N or ORF1AB Ct < 30 and S gene undetermined. The data were aggregated on a week and state level for surveillance purposes.

## Quantification and statistical analysis

### Airport catchment model

The Huff model is a probabilistic approach which is traditionally used to determine the probability that a given population will go to a specific service location (Huff, 1963, 2003). Recently it has been applied to determine airport catchment areas, i.e. the location someone will go after arriving at a certain airport (Huber et al., 2021). This approach incorporates the distance from an airport to surrounding counties (or other geographical units such as census tract) and the attractiveness of that airport. The Huff model is represented as (Huff, 2003):

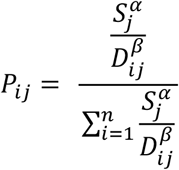

Where P_ij_ represents the probability that individuals that arrive at airport j will go to county i, S_j_ is a measure of attractiveness for airport j, D_ij_ is the distance from county i to airport j,*α*is an airport attractiveness exponent, and *β*is the distance decay exponent. As a proxy for S_j_ we use the number of passengers arriving at airport j from the United Kingdom in December 2020. Based on previous analysis optimizing the Huff model for airport catchment models, we set *α*to 1 and *β* to 2.

Huber and Rinner recommended the use of a distance cut-off so that the catchment area represents a reasonable maximum distance that a person would be willing to travel from an airport (Huber and Rinner, 2020). We selected a distance cut-off of 200km, because this represents a conservative estimate of which counties surrounding each airport are most likely for people to travel to once they deplane (Huber et al., 2021). Using flight data from OAG, we selected airports in the US that received at least one percent of the total passengers from the UK in December 2020. We utilized a Huff model for each of these airports, and combined these results to estimate the approximate number of people in the counties surrounding the airports that traveled from the UK. These results are displayed in **Figures 1A-B**.

### Phylogenetic analysis

To perform phylogenetic analyses, we initially build a dataset containing only B.1.1.7 genomes (genomes, n=101,079) based on data available up to February 26, 2021 on GISAID (gisaid.org). Since the proportion of B.1.1.7 cases is not known for most countries, we subsampled this set of genomes according to the proportion of overall COVID-19 cases reported per epidemiological week in each country, using data from the Johns Hopkins University, Center for Systems Science and Engineering (CSSE) (github.com/CSSEGISandData/COVID-19). The subsampling was performed using the pipeline ‘subsampler’ (github.com/andersonbrito/subsampler), which selected available genomes simulating a scenario of 0.1% of sequenced cases per epiweek, per country. This allowed us to obtain a dataset with 8,864 B.1.1.7 genomes from 59 countries (7,589 international, and 1,275 from the US, all with coverage above 70%), representative of the COVID-19 burden revealed by the epidemiological time series data from each country. As part of this dataset, there were 770 genomes that we sequenced from 20 US states from 2020-12-19 to 2021-02-14, provided by the CDC (568), New York State Department of Health (41), University of Michigan (45), and Yale University (116). The B.1.1.7 samples sequenced by the CDC were generated for public health surveillance, and we received direct permission to download the sequences from GISAID for primary analysis. The final dataset was composed by 8,864B.1.1.7 genomes, and one P.1 genome to root the tree, serving as an outgroup (Brazil/AM-20842882CA/2020). The complete list of genomes, with author acknowledgements, can be found in **Data S2**.

Using an augur pipeline (Hadfield et al., 2018), we performed multiple sequence alignment (MSA) using MAFFT (Katoh and Standley, 2013), and the 5’ and 3’ ends of the MSA were masked alongside other problematic sites (Maio et al., 2020) using a script provided with the pipeline. A quick maximum likelihood analysis was performed using IQ-Tree (Minh et al., 2020) under a GTR nucleotide substitution model. Inference of divergence times and reconstruction of ancestral states were performed using TreeTime 0.8.0 (Sagulenko et al., 2018). This preliminary analysis aimed at determining the placement of the US B.1.1.7 genomes with respect to international samples (**Figure S2**), and at removing major molecular clock outliers (n=34) deviating more than 4 interquartile ranges from the root-to-tip regression line. This phylogeny was then used for identifying large clades containing only genomes of European or Global origin, which were then individually pruned down to contain only three representatives per clade. This procedure dramatically decreased the total number of genomes in the dataset from 8,864 to 1,913, while still keeping the same overall topology. The smaller dataset (1,913 B.1.1.7 genomes) was then run through the same pipeline as described above, but with 1,000 UFBoot replicates during tree inference using IQ-Tree (Minh et al., 2013). This tree served as input for a root-to-tip analysis using TempEst (Rambaut et al., 2016), where 5 outliers with residual above ± 0.0002 subs/site were removed (**Figure S3**). With a clean set of 1,908 genomes we proceeded with the inference of the final time-scaled tree using TreeTime (Sagulenko et al., 2018).

Finally, using the time-scaled maximum-likelihood tree as a fixed topology, we performed Bayesian inference of ancestral states (discrete phylogeographic reconstruction) using BEAST v.1.10 (Suchard et al., 2018), for 15×10^6^ generations, sampling every 1,000 generations, which led to MCMC convergence and good mixing, with all parameters showing ESS > 200 when assessed using Tracer 1.7 (Rambaut et al., 2018). After discarding 10% of the sampled trees as burn-in, we used TreeAnnotator to obtain the final tree, identical to the TreeTime output, but with ancestral states inferred by BEAST. We visualized this time-scaled maximum-likelihood phylogeny using auspice (Hadfield et al., 2018), which can be found on our custom nextstrain page: nextstrain.org/community/grubaughlab/CT-SARS-CoV-2/paper5. We combined bootstrap support and Bayesian inference of ancestral states with our time-scaled maximum-likelihood tree shown in **Figure 3** using baltic (github.com/evogytis/baltic). To identify independent, international introductions of B.1.1.7 in the US, we selected well supported clades (UFBoot > 70; MRCA discrete state probability > 0.7) with 3 or more taxa and represented these clades in **Figure 3B** using the “exploded tree view” from baltic (phylo-baltic.github.io/baltic-gallery/basic-exploded-tree-flu/), to highlight changes in ancestral state (International origin > USA). The clades shown in **Figures 3C-G** are zoom highlights from the large tree shown in **Figure 3A**.

To externally validate the time scale and tMRCA estimates from TreeTime, we utilized the dataset used in the inference of the Bayesian MCC tree generated by Washington etl al (github.com/andersen-lab/paper_2021_early-b117-usa) to reconstruct a similar time-resolved phylogeny using TreeTime for comparison. We analyzed 4 separate US clades using both methods and found that they produced similar mean tMRCAs and overlapping temporal distributions (**Figure S4**).

Finally, in order to compare the discrete ancestral state reconstructions fully inferred using TreeTime 0.8.0 with inferences done using BEAST v.1.10, we plotted the results from TimeTree using the same approach used to generate **Figure 3**, to create **Figure S5**, to highlight that results obtained through maximum-likelihood analysis (using TreeTime) is equivalent to those obtained in more computing intensive Bayesian analysis (using BEAST). The results presented in **Figure 3** use BEAST for the discrete ancestral state reconstructions.

### Projection of SGTF data

A logistic growth model was fit to the weekly SGTF quantification for each state using the glm() function from the stats package in R. Code is available on github.com/grubaughlab/paper_2021_B117-US.

## Resource Availability

### Lead contact

Further information and requests for data, resources, and reagents should be directed to and will be fulfilled by the Lead Contact, Nathan D. Grubaugh.

### Materials availability

#### Data and code availability

Data used to produce all of the figures are included in **Data S1** and **Data S2** and on github.com/grubaughlab/paper_2021_B117-US along with all code used for analyses. The subsampling pipeline can be found on github.com/andersonbrito/subsampler. Genomic data are available on GISAID (see **Data S2** for accession numbers). The air passenger data used in this study are proprietary and were purchased from OAG Aviation Worldwide Ltd. These data were used under the United States Centers for Disease Control and Prevention license for the current study and so are not publicly available. The authors are available to share the air passenger data upon reasonable request and with the permission of OAG Aviation Worldwide Ltd.

### Supplemental Information Description

**Data S1**. Data for Figures 1-4.

**Data S2**. List of SARS-CoV-2 sequences used in this study and author acknowledgements.

## Supporting information

DataS1

DataS2

## Data Availability

Data used to produce all of the figures are included in Data S1 and Data S2 and on github.com/grubaughlab/paper_2021_B117-US along with all code used for analyses. The subsampling pipeline can be found on github.com/andersonbrito/subsampler. Genomic data are available on GISAID (see Data S2 for accession numbers). The air passenger data used in this study are proprietary and were purchased from OAG Aviation Worldwide Ltd. These data were used under the United States Centers for Disease Control and Prevention license for the current study and so are not publicly available. The authors are available to share the air passenger data upon reasonable request and with the permission of OAG Aviation Worldwide Ltd.

## Acknowledgments

We thank L. Cong and P. Ruggiero for support, K. Gangavarapu and K. Anderson for making their phylogenetic analysis openly available, the frontline and essential workers for their continued service during the pandemic, and our friends and family - particularly V. Parsons, P. Jack, and S. Taylor - for their support. This work was funded by CTSA Grant Number TL1 TR001864 (T.A. and M.E.P.), Fast Grant from Emergent Ventures at the Mercatus Center at George Mason University (N.D.G.), CDC Contract # 75D30120C09570 (N.D.G.), CDC Contract # 75D30120C09870 (A.S.L.). Initial funding for sequencing at the Wadsworth Center was provided by the New York Community Trust. The findings and conclusions in this report are those of the authors and do not necessarily represent the official position of the Centers for Disease Control and Prevention. Use of trade names is for identification only and does not imply endorsement by the Centers for Disease Control and Prevention.

## Author Contributions

Conceptualization,T.A., E. L-N., A.F.B., A.L.V., J.R., M.J.M., J.R.F, C.E.M, A.S.L, K.S.G, D.R.M., N.D.G.; Investigation,T.A., E.L-N., A.F.B., A.L.V., J.R., M.J.M., M.E.P., M.I.B., A.E.W., C.C.K., S.D., G.K., C.E.M., J.W., M.L.L., A.G., S.M.M., H.R., M.C., L.W., P.V., A.C., K.A.F., D.R.P.; Writing - Original Draft, T.A., E.L-N., A.F.B., J.F., N.D.G; Writing - Review & Editing, T.A., E. L-N., A.F.B., A.L.V., J.R., M.J.M., M.E.P., M.I.B., A.E.W., C.B.F.V., C.C.K., S.D., A.R., J.P.K.,M.S., J.P., E.S., W.J.F., G.K., J.M., J.T.D., M.N., N.B., J.W., C.L., P.H., A.M., R.D., J.R., S.M.B., A.G., S.M.M., S.M., C.N., E.L., H.R., M.C., L.W., P.V., A.C., K.A.F., D.R.P., M.L.L., P.C., J.R.F, C.E.M, A.S.L, K.S.G, D.R.M., N.D.G.

## Supplemental Information

**Table S1. Related to Figure 1.**
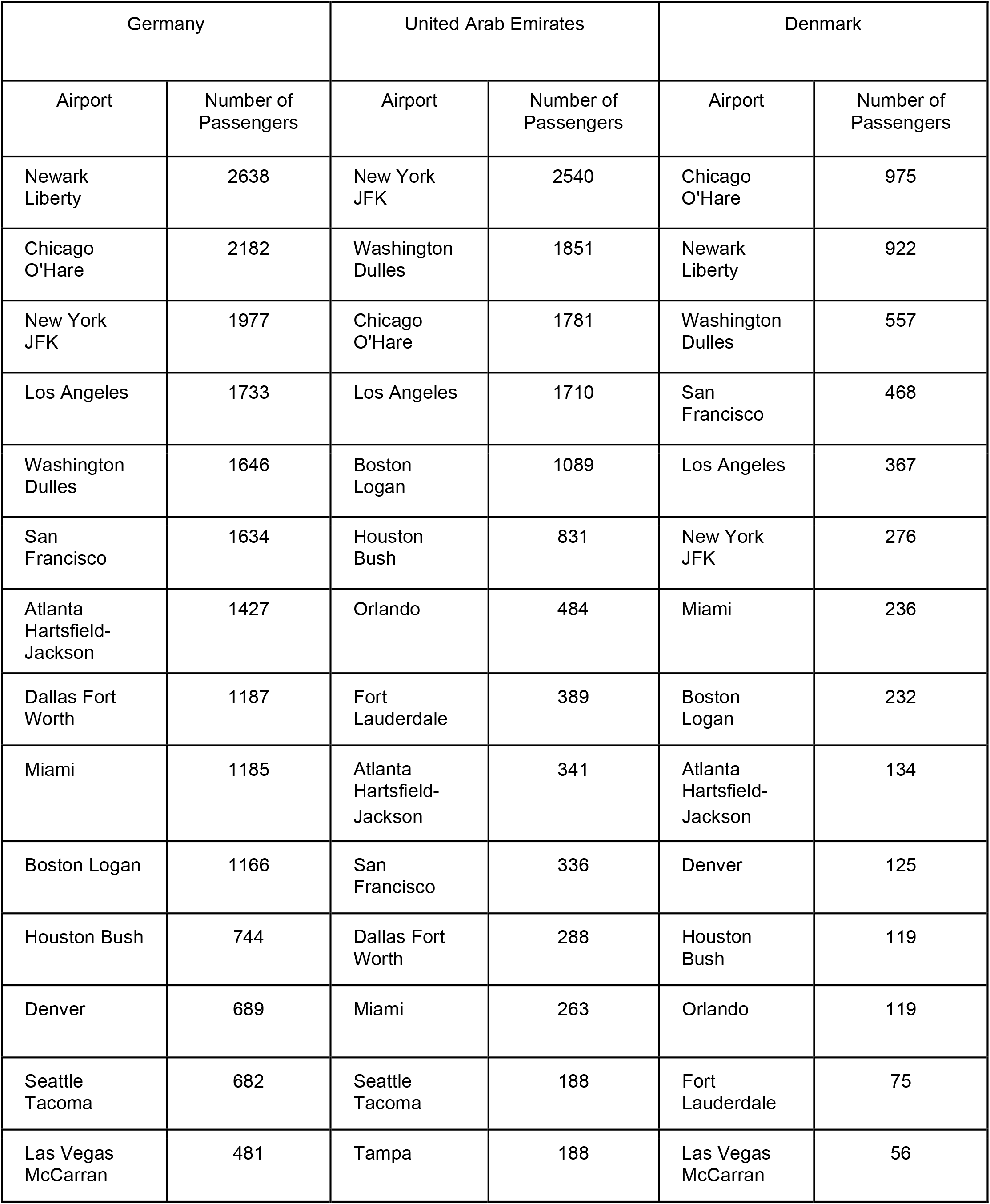
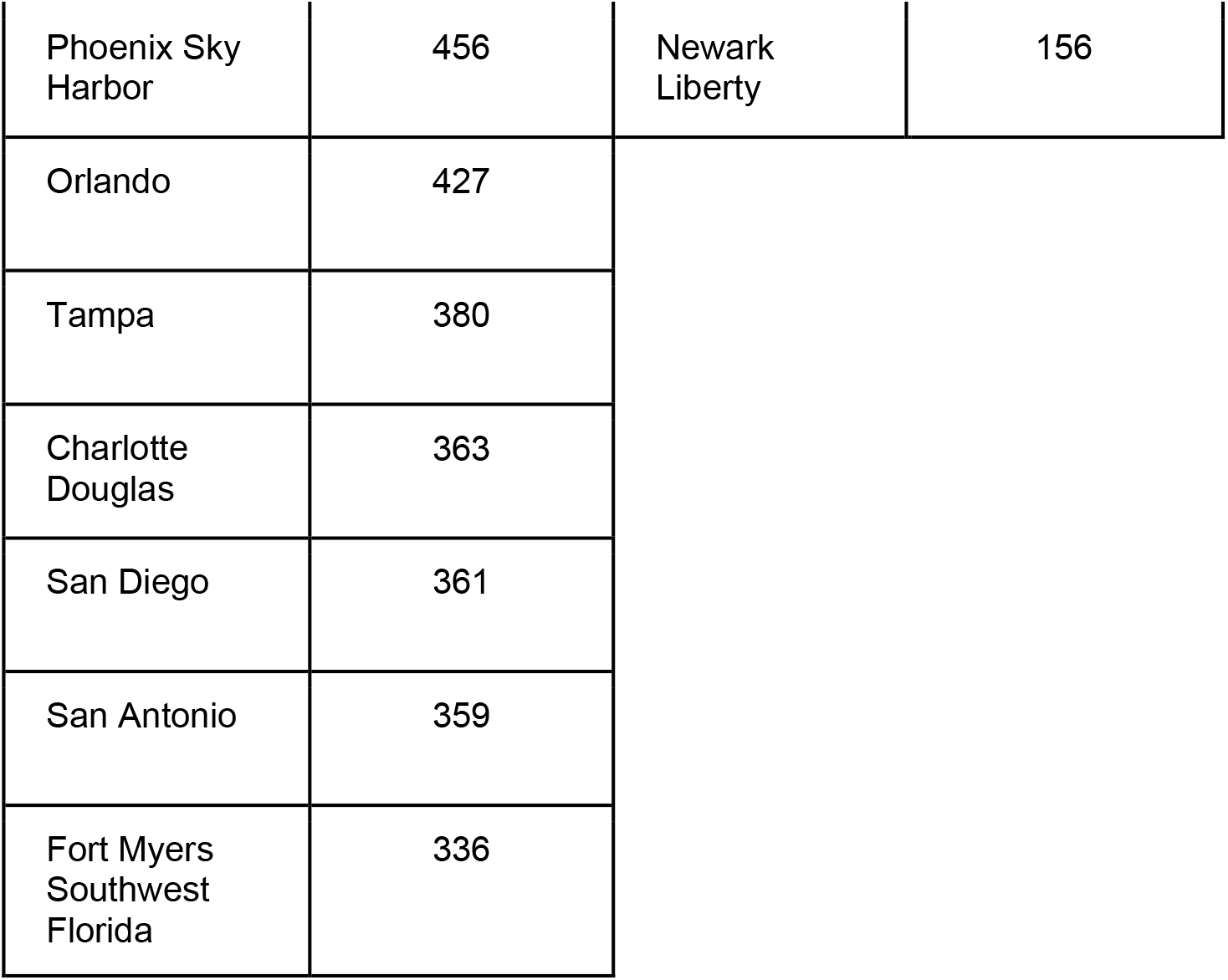
Number of passengers from Germany, United Arab Emirates, and Denmark inbound to top United States airports (defined as airports with at least 1% of total travel from each country to the US) in December 2020.

| Germany |  | United Arab Emirates |  | Denmark |  |
| --- | --- | --- | --- | --- | --- |
| Airport | Number of Passengers | Airport | Number of Passengers | Airport | Number of Passengers |
| Newark Liberty | 2638 | New York JFK | 2540 | Chicago O'Hare | 975 |
| Chicago O'Hare | 2182 | Washington Dulles | 1851 | Newark Liberty | 922 |
| New York JFK | 1977 | Chicago O'Hare | 1781 | Washington Dulles | 557 |
| Los Angeles | 1733 | Los Angeles | 1710 | San Francisco | 468 |
| Washington Dulles | 1646 | Boston Logan | 1089 | Los Angeles | 367 |
| San Francisco | 1634 | Houston Bush | 831 | New York JFK | 276 |
| Atlanta Hartsfield-Jackson | 1427 | Orlando | 484 | Miami | 236 |
| Dallas Fort Worth | 1187 | Fort Lauderdale | 389 | Boston Logan | 232 |
| Miami | 1185 | Atlanta Hartsfield-Jackson | 341 | Atlanta Hartsfield-Jackson | 134 |
| Boston Logan | 1166 | San Francisco | 336 | Denver | 125 |
| Houston Bush | 744 | Dallas Fort Worth | 288 | Houston Bush | 119 |
| Denver | 689 | Miami | 263 | Orlando | 119 |
| Seattle Tacoma | 682 | Seattle Tacoma | 188 | Fort Lauderdale | 75 |
| Las Vegas McCarran | 481 | Tampa | 188 | Las Vegas McCarran | 56 |

|  |  |  |  |
| --- | --- | --- | --- |
| Phoenix Sky Harbor | 456 | Newark Liberty | 156 |
| Orlando | 427 |  |  |
| Tampa | 380 |  |  |
| Charlotte Douglas | 363 |  |  |
| San Diego | 361 |  |  |
| San Antonio | 359 |  |  |
| Fort Myers Southwest Florida | 336 |  |  |

**Figure S1. Related to Figure 2.**
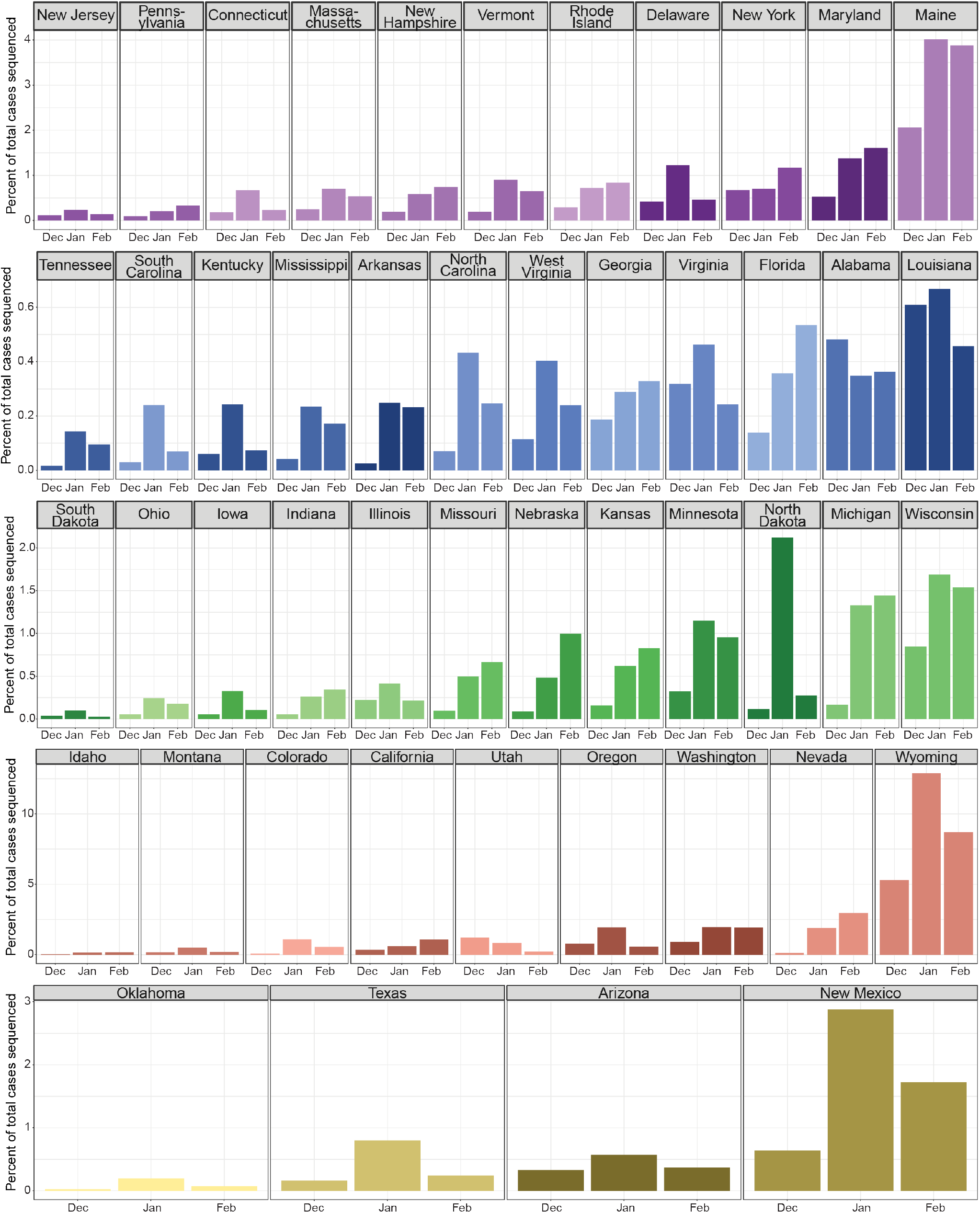
The percentage of total COVID-19 cases which were sequenced in December 2020 (Dec), January 2021 (Jan), and February 2021 (Feb) in each state of the continental US. Color legend is the same as in Figure 2A.

**Figure S2. Related to Figure 3.**
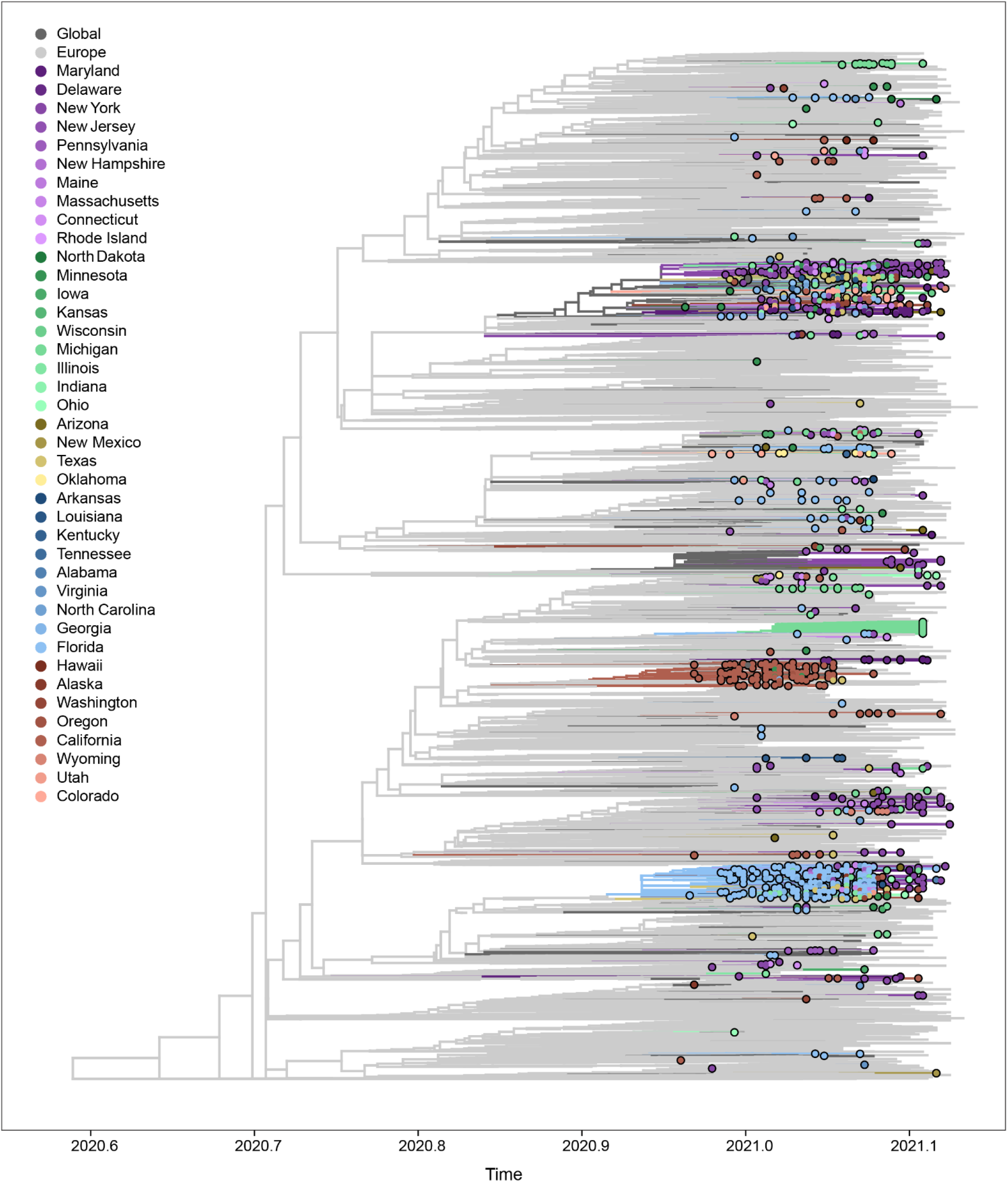
Maximum likelihood phylogeny of B.1.1.7, including 8,829 representative genomes from the US, Europe, other global locations. Phylogenetic inference was performed using IQ-Tree 1.6.12, with time scale and discrete state reconstruction inferred using TreeTime 0.8.0, and data visualization using baltic 0.1.5. US B.1.1.7 genomes are highlighted with circles at the tips, while international genomes are only represented as branches. The tree was rooted using a P.1 genome (Brazil/AM-20842882CA/2020) as an outgroup (not shown in this plot). This larger dataset was used to further subsample the genomes, removing redundant B.1.1.7 clades containing only genomes of international origin. From this phylogeny we created a succinct dataset containing 1,908 shown in **Figure 3**.

**Figure S3. Related to Figure 3.**
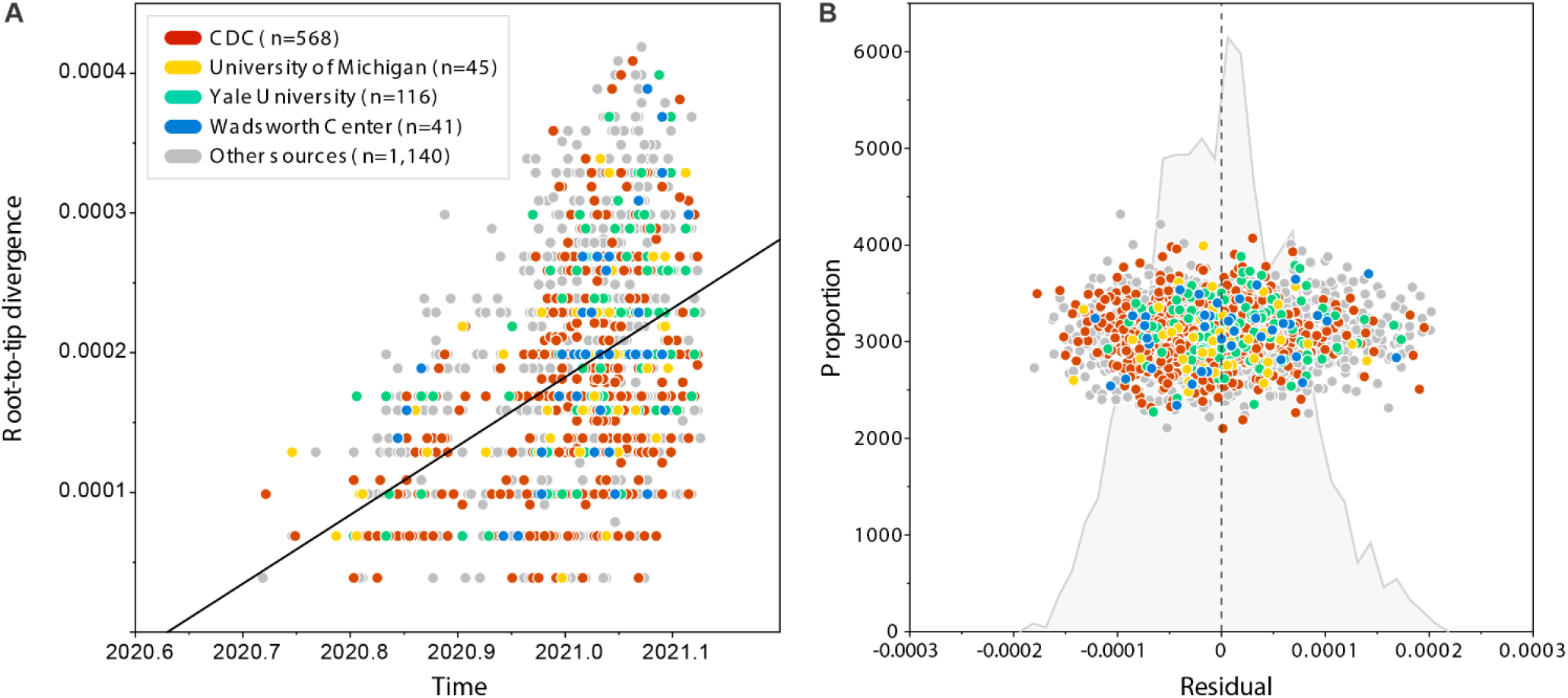
Root-to-tip analysis of 1,908 B.1.1.7 genomes used to obtain the phylogenetic results shown in Figure 3. A. Correlation between genetic divergence (subs/site) and time. Samples generated in this study are highlighted with colours, while background international genomes are shown on grey. B. Distribution of genetic divergence residuals of genomes shown in (A). Any outliers with residuals above ± 0.0002 subs/site were removed from downstream analyses.

**Figure S4. Related to Figure 3.**
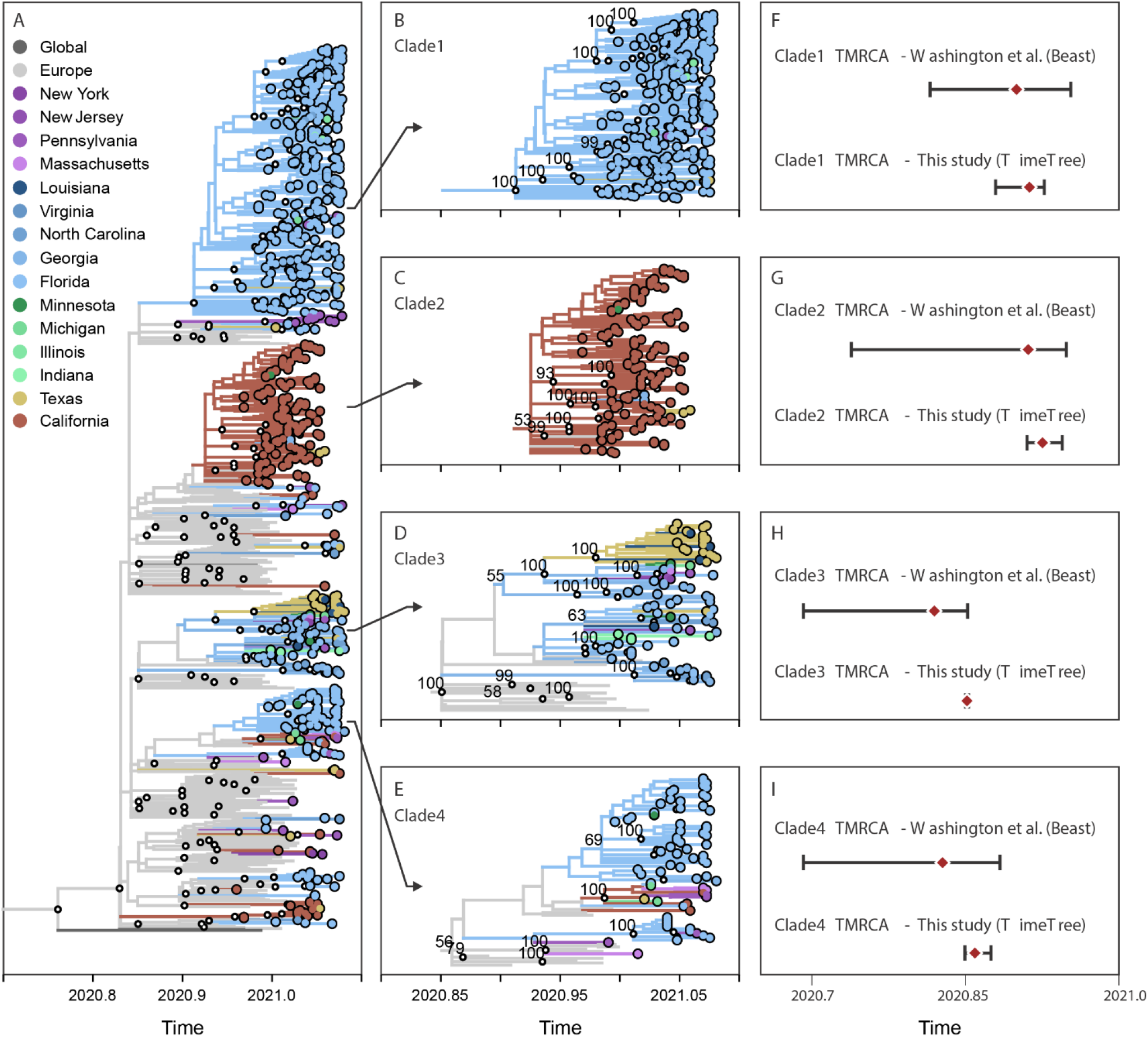
Re-analysis of phylogenetic results by Washignton et al. (Washington et al., 2021). **A**. Tree topology and bootstrap values (UFBoot > 70 represented by small black circles at the nodes) obtained using IQ-Tree 1.6.12, with time scale and discrete state reconstruction inferred by TreeTime 0.8.0, and data integration and visualization using baltic 0.1.5. Like the original analysis, the tree was rooted using the genome Wuhan/Hu-1/2019 as an outgroup (not shown in this plot). **B-E**. Four clades of US B.1.1.7 genomes selected for comparison of time scales. **F-I**. Comparison of median and confidence intervals of tMRCAs obtained in the original study by Washignton et al. (at the top) and in our analysis using TreeTime (at the bottom of each panel).

**Figure S5. Related to Figure 3.**
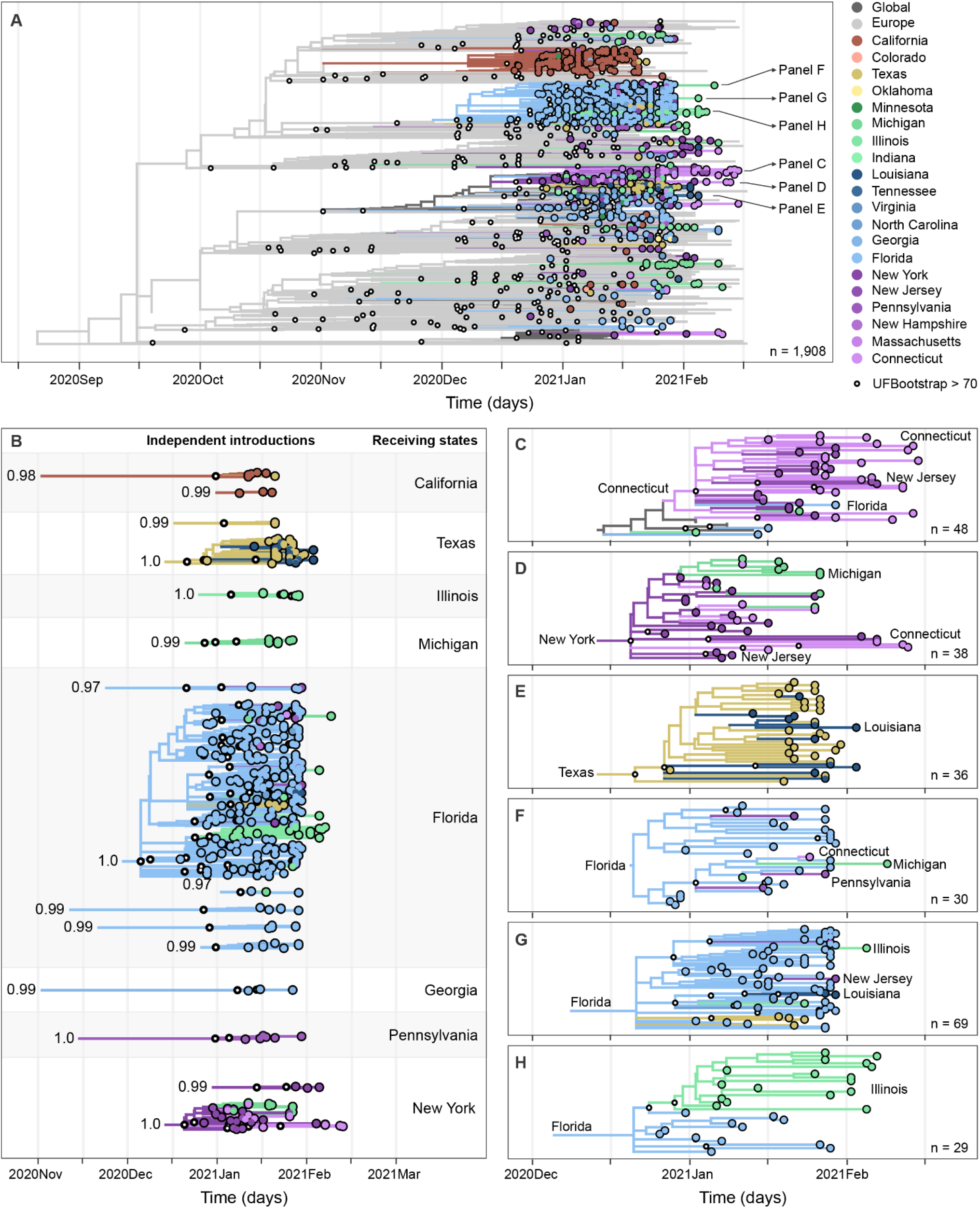
Results obtained using TreeTime only, plotted using the same approach used for Figure 3, showing multiple introductions, domestic spread, and community transmission of B.1.1.7 SARS-CoV-2 in the United States. **A**. Maximum likelihood phylogeny of B.1.1.7, including 1,908 representative genomes from the US, Europe, other global locations. Tree topology and bootstrap values obtained using IQ-Tree 1.6.12, with time scale and discrete state reconstruction inferred by TreeTime 0.8.0, and data integration and visualization using baltic 0.1.5. The tree was rooted using a P.1 genome (Brazil/AM-20842882CA/2020) as an outgroup (not shown in this plot). **B**. Exploded tree layout, highlighting clades with 3 or more taxa, UFBoot > 70 (small circles), and US ancestral state probability at MRCA > 0.7 (values at the root), representing independent international introductions of B.1.1.7 into distinct regions of the US, based on the same phylogenetic tree shown in (**A**). A list of international transitions to the US can be found in **Data S1. C-HE**.Time-informed maximum likelihood phylogeny of distinct B.1.1.7 clades showing instances of intra-region (**C, D, E, G**) and inter-region (**D, H**) domestic spread. (**C**,**E**) and/or community transmission within New York (**C**), Connecticut (**C**), Michigan (**C**,**D**), and Illinois (**E**). The list of SARS-CoV-2 sequences used in this study and author acknowledgements can be found in **Data S2**. Supporting phylogenetic analysis can be found in **Figures S2-5**. For comparison, an interactive phylogenetic tree, inferred using IQ-Tree and TreeTime only, can be accessed from our custom Nextstrain build: **Table S1. Related to Figure 1**. Number of passengers from Germany, United Arab Emirates, and Denmark inbound to top United States airports (defined as airports with at least 1% of total travel from each country to the US) in December 2020. https://nextstrain.org/community/grubaughlab/CT-SARS-CoV-2/paper5

## References

Becker, R.A., Wilks, A.R., Brownrigg, R., Minka, T.P., and Deckmyn, A. (2018). maps: Draw Geographical Maps. R package version 3.3. 0.

Borges, V., Sousa, C., Menezes, L., A, M.G., Picão, M., Almeida, J.P., Vieita, M., Santos, R., Silva, A.R., Costa, M., et al. (2021). Tracking SARS-CoV-2 VOC 202012/01 (lineage B.1.1.7) dissemination in Portugal: insights from nationwide RT-PCR Spike gene drop out data.

CDC (2021a). US COVID-19 Cases Caused by Variants.

CDC (2021b). Genomic Surveillance for SARS-CoV-2.

Davies, N.G., Abbott, S., Barnard, R.C., Jarvis, C.I., Kucharski, A.J., Munday, J.D., Pearson, C.A.B., Russell, T.W., Tully, D.C., Washburne, A.D., et al. (2021). Estimated transmissibility and impact of SARS-CoV-2 lineage B.1.1.7 in England. Science.

Dellicour, S., Durkin, K., Hong, S.L., Vanmechelen, B., Martí-Carreras, J., Gill, M.S., Meex, C., Bontems, S., André, E., Gilbert, M., et al. (2020). A Phylodynamic Workflow to Rapidly Gain Insights into the Dispersal History and Dynamics of SARS-CoV-2 Lineages. Mol. Biol. Evol.

Faria, N.R., Claro, I.M., Candido, D., Moyses Franco, L.A., Andrade, P.S., Coletti, T.M., Silva, C.A.M., Sales, F.C., Manuli, E.R., Aguiar, R.S., et al. (2021). Genomic characterisation of an emergent SARS-CoV-2 lineage in Manaus: preliminary findings. Virological.

Fauver, J.R., Petrone, M.E., Hodcroft, E.B., Shioda, K., Ehrlich, H.Y., Watts, A.G., Vogels, C.B.F., Brito, A.F., Alpert, T., Muyombwe, A., et al. (2020). Coast-to-Coast Spread of SARS-CoV-2 during the Early Epidemic in the United States. Cell 181, 990–996.e5.

Galloway, S.E., Paul, P., MacCannell, D.R., Johansson, M.A., Brooks, J.T., MacNeil, A., Slayton, R.B., Tong, S., Silk, B.J., Armstrong, G.L., et al. (2021). Emergence of SARS-CoV-2 B.1.1.7 Lineage -United States, December 29, 2020-January 12, 2021. MMWR Morb. Mortal. Wkly. Rep. 70, 95–99.

Gonzalez-Reiche, A.S., Hernandez, M.M., Sullivan, M.J., Ciferri, B., Alshammary, H., Obla, A., Fabre, S., Kleiner, G., Polanco, J., Khan, Z., et al. (2020). Introductions and early spread of SARS-CoV-2 in the New York City area. Science 369, 297–301.

Grubaugh, N.D., Hodcroft, E.B., Fauver, J.R., Phelan, A.L., and Cevik, M. (2021). Public health actions to control new SARS-CoV-2 variants. Cell 0.

Hadfield, J., Megill, C., Bell, S.M., Huddleston, J., Potter, B., Callender, C., Sagulenko, P., Bedford, T., and Neher, R.A. (2018). Nextstrain: real-time tracking of pathogen evolution. Bioinformatics 34, 4121– 4123.

Huber, C., and Rinner, C. (2020). Market Area Delineation for Airports to Predict the Spread of Infectious Disease. In Geospatial Technologies for Local and Regional Development, (Springer International Publishing), pp. 263–289.

Huber, C., Watts, A., Grills, A., Yong, J.H.E., Morrison, S., Bowden, S., Tuite, A., Nelson, B., Cetron, M., and Khan, K. (2021). Modelling airport catchment areas to anticipate the spread of infectious diseases across land and air travel. Spat. Spatiotemporal Epidemiol. 36, 100380.

Huff, D.L. (1963). A Probabilistic Analysis of Shopping Center Trade Areas. Land Econ. 39, 81–90.

Huff, D.L. (2003). Parameter estimation in the Huff model. Esri, ArcUser 34–36.

Katoh, K., and Standley, D.M. (2013). MAFFT multiple sequence alignment software version 7: improvements in performance and usability. Mol. Biol. Evol. 30, 772–780.

Kemp, S.A., Collier, D.A., Datir, R., Ferreira, I.A., Gayed, S., Jahun, A., Hosmillo, M., Rees-Spear, C., Mlcochova, P., Lumb, I.U., et al. (2020). Neutralising antibodies drive Spike mediated SARS-CoV-2 evasion (medRxiv).

Larsen, B.B., and Worobey, M. (2020). Identification of a novel SARS-CoV-2 Spike 69-70 deletion lineage circulating in the United States.

Lauring, A.S., and Hodcroft, E.B. (2021). Genetic Variants of SARS-CoV-2-What Do They Mean? JAMA.

Lemey, P., Rambaut, A., Drummond, A.J., and Suchard, M.A. (2009). Bayesian phylogeography finds its roots. PLoS Comput. Biol. 5, e1000520.

Loman, N.J., Quick, J., and Simpson, J.T. (2015). A complete bacterial genome assembled de novo using only nanopore sequencing data. Nat. Methods 12, 733–735.

Maio, D., Walker, C., Borges, R., Weilguny, L., Slodkowicz, G., Goldman, N., and Nicola (2020). Masking strategies for SARS-CoV-2 alignments.

Maurano, M.T., Ramaswami, S., Zappile, P., Dimartino, D., Boytard, L., Ribeiro-Dos-Santos, A.M., Vulpescu, N.A., Westby, G., Shen, G., Feng, X., et al. (2020). Sequencing identifies multiple early introductions of SARS-CoV-2 to the New York City region. Genome Res. 30, 1781–1788.

Minh, B.Q., Nguyen, M.A.T., and von Haeseler, A. (2013). Ultrafast approximation for phylogenetic bootstrap. Mol. Biol. Evol. 30, 1188–1195.

Minh, B.Q., Schmidt, H.A., Chernomor, O., Schrempf, D., Woodhams, M.D., von Haeseler, A., and Lanfear, R. (2020). IQ-TREE 2: New Models and Efficient Methods for Phylogenetic Inference in the Genomic Era. Mol. Biol. Evol. 37, 1530–1534.

Moreno, G., Braun, K., Larsen, B.B., Alpert, T., Worobey, M., Grubaugh, N., Friedrich, T., O’Connor, D., Fauver, J., and Brito, A. (2021). Detection of non-B.1.1.7 spike Δ69/70 sequences (B.1.375) in the United States.

PHE (2020). Investigation of novel SARS-CoV-2 variant, Technical Briefing 3. Public Health England 3.

Quick, J. (2020). nCoV-2019 sequencing protocol v3.

Quick, J., Grubaugh, N.D., Pullan, S.T., Claro, I.M., Smith, A.D., Gangavarapu, K., Oliveira, G., Robles-Sikisaka, R., Rogers, T.F., Beutler, N.A., et al. (2017). Multiplex PCR method for MinION and Illumina sequencing of Zika and other virus genomes directly from clinical samples. Nat. Protoc. 12, 1261–1276.

Rambaut, A., Lam, T.T., Max Carvalho, L., and Pybus, O.G. (2016). Exploring the temporal structure of heterochronous sequences using TempEst (formerly Path-O-Gen). Virus Evol 2, vew007.

Rambaut, A., Drummond, A.J., Xie, D., Baele, G., and Suchard, M.A. (2018). Posterior Summarization in Bayesian Phylogenetics Using Tracer 1.7. Syst. Biol. 67, 901–904.

Rambaut, A., Loman, N., Pybus, O., Barclay, W., Barrett, J., Carabelli, A., Connor, T., Peacock, T., Robertson, D.L., and Volz, E. (2020). Preliminary genomic characterisation of an emergent SARS-CoV-2 lineage in the UK defined by a novel set of spike mutations. Virological.

Sagulenko, P., Puller, V., and Neher, R.A. (2018). TreeTime: Maximum-likelihood phylodynamic analysis. Virus Evol 4, vex042.

Starr, T.N., Greaney, A.J., Hilton, S.K., Ellis, D., Crawford, K.H.D., Dingens, A.S., Navarro, M.J., Bowen, J.E., Tortorici, M.A., Walls, A.C., et al. (2020). Deep Mutational Scanning of SARS-CoV-2 Receptor Binding Domain Reveals Constraints on Folding and ACE2 Binding. Cell 182, 1295–1310.e20.

Suchard, M.A., Lemey, P., Baele, G., Ayres, D.L., Drummond, A.J., and Rambaut, A. (2018). Bayesian phylogenetic and phylodynamic data integration using BEAST 1.10. Virus Evol 4, vey016.

Tegally, H., Wilkinson, E., Giovanetti, M., Iranzadeh, A., Fonseca, V., Giandhari, J., Doolabh, D., Pillay, S., San, E.J., Msomi, N., et al. (2020). Emergence and rapid spread of a new severe acute respiratory syndrome-related coronavirus 2 (SARS-CoV-2) lineage with multiple spike mutations in South Africa (medRxiv).

Vavrek, D., Speroni, L., Curnow, K.J., Oberholzer, M., Moeder, V., and Febbo, P.G. (2021). Genomic surveillance at scale is required to detect newly emerging strains at an early timepoint (medRxiv).

Vogels, C.B.F., Breban, M., Alpert, T., Petrone, M.E., Watkins, A.E., Hodcroft, E.B., Mason, C.E., Khullar, G., Metti, J., Dudley, J.T., et al. (2021). PCR assay to enhance global surveillance for SARS-CoV-2 variants of concern (medRxiv).

Volz, E., Mishra, S., Chand, M., Barrett, J.C., Johnson, R., Geidelberg, L., Hinsley, W.R., Laydon, D.J., Dabrera, G., O’Toole, Á., et al. (2021). Transmission of SARS-CoV-2 Lineage B.1.1.7 in England: Insights from linking epidemiological and genetic data (medRxiv).

Washington, N.L., Gangavarapu, K., Zeller, M., Bolze, A., Cirulli, E.T., Schiabor Barrett, K.M., Larsen, B.B., Anderson, C., White, S., Cassens, T., et al. (2021). Genomic epidemiology identifies emergence and rapid transmission of SARS-CoV-2 B.1.1.7 in the United States. medRxiv 2021.02.06.21251159.

Wickham, H. (2016). GGPLOT2: Elegant Graphics for Data Analysis 2016 Springer-Verlag, New York.

Wise, J. (2021). Covid-19: The E484K mutation and the risks it poses. BMJ 372, 359.

Zimmer, C., and Pietsch, B. (2020). First U.S. Case of Highly Contagious Coronavirus Variant Is Found in Colorado. The New York Times.

Huber C, Rinner C. (2020). Market Area Delineation for Airports to Predict the Spread of Infectious Disease. In: Kyriakidis P, Hadjimitsis D, Skarlatos D, Mansourian A, editors. Geospatial Technologies for Local and Regional Development. Springer; p. 263–89.

Lamstein, A., Johnson, B., and Trulia, Inc. (2020). choroplethr: Simplify the Creation of Choropleth Maps in R. R package version 3.7.0.

